# A Pilot Randomized Controlled Trial of *de novo* Belatacept-Based Immunosuppression in Lung Transplantation

**DOI:** 10.1101/2021.11.12.21265985

**Authors:** Howard J. Huang, Kenneth Schechtman, Medhat Askar, Cory Bernadt, Brigitte Mittler, Peter Dore, Chad Witt, Derek Byers, Rodrigo Vazquez-Guillamet, Laura Halverson, Ruben Nava, Varun Puri, Andrew Gelman, Daniel Kreisel, Ramsey R. Hachem

**Affiliations:** Houston Methodist Hospital; Division of Biostatistics, Washington University in St. Louis; Department of Pathology and Laboratory Medicine, Texas A & M College of Medicine; Department of Pathology and Immunology, Washington University in St. Louis; Division of Pulmonary and Critical Care, Washington University in St. Louis; Division of Cardiothoracic Surgery, Washington University in St. Louis

## Abstract

The development of donor-specific antibodies (DSA) after lung transplantation is common and results in adverse outcomes. In kidney transplantation, Belatacept has been associated with a lower incidence of DSA, but experience with Belatacept in lung transplantation is limited. We conducted a 2-center pilot randomized controlled trial of *de novo* immunosuppression with Belatacept after lung transplantation to assess the feasibility of conducting a pivotal trial. Twenty-seven participants were randomized to Control (Tacrolimus, Mycophenolate Mofetil, and prednisone, n = 14) or Belatacept-based immunosuppression (Tacrolimus, Belatacept, and prednisone until day 89 followed by Belatacept, Mycophenolate Mofetil, and prednisone, n = 13). All participants were treated with rabbit anti-thymocyte globulin for induction immunosuppression. We permanently stopped randomization and treatment with Belatacept after 3 participants in the Belatacept arm died compared to none in the Control arm. Subsequently, 2 additional participants in the Belatacept arm died for a total of 5 deaths compared to none in the Control arm (log rank p = 0.016). We did not detect a significant difference in DSA development, acute cellular rejection, or infection between the 2 groups. We conclude that this investigational regimen using Belatacept after lung transplantation is associated with significantly increased mortality.

## INTRODUCTION

Lung transplantation is the ultimate treatment for patients with advanced lung disease, and approximately 4,500 patients undergo lung transplantation annually worldwide (1). The median survival after lung transplantation is 6 years, and the leading cause of death beyond the first year is chronic lung allograft dysfunction (CLAD) (1). Although the exact pathogenesis of CLAD is unknown, clinical risk factors have been identified. These include acute cellular rejection (ACR), lymphocytic bronchiolitis (LB), antibody-mediated rejection (AMR), and the development of donor-specific antibodies (DSA) to mismatched human leukocyte antigens (HLA) (2-7). The development of DSA is an independent risk factor for ACR, LB, and DSA directly cause AMR (4, 5). Furthermore, the development of DSA has been identified as an independent risk factor for death (6).

Cellular immune responses were recognized early in the history of transplantation as the primary barrier to organ acceptance. Therapeutic suppression of T-cell function and proliferation made organ transplantation clinically feasible. However, rejection remains a persistent obstacle to better long-term outcomes, especially among lung transplant recipients. Furthermore, a significant proportion of lung transplant recipients treated with conventional immunosuppression develop DSA early after transplantation. In a prospective multicenter observational study, 36% of lung recipients developed DSA within 120 days of transplantation (8). In other studies, the incidence of DSA in the first year after transplantation has been as high as 61% (6). This illustrates that conventional immunosuppression does not sufficiently suppress alloantibody production or its effects on the allograft.

T-cell activation requires 2 synergistic signals (9-11). The first is recognition by the T-cell receptor of antigen bound by MHC molecules on antigen presenting cells. The second signal comprises engagement of co-stimulatory molecules CD28 and CTLA4 expressed on T-cells to their cognate ligands CD80 and CD86 expressed on antigen presenting cells to modulate T-cell activity. In the absence of co-stimulatory signals, T-cells become anergic or undergo apoptosis (12, 13). Belatacept, a CTLA4-Ig fusion protein that binds CD80 and CD86 thereby blocking CD28 co-stimulatory signals, is approved for the prevention of kidney transplant rejection. In a multicenter randomized controlled trial (RCT), kidney recipients treated with Belatacept had significantly better patient and allograft survival than those treated with Cyclosporine, and there was no difference in the incidence of serious adverse events (SAE) or serious infections between the groups (14). Furthermore, recipients treated with Belatacept were significantly less likely to develop DSA (14). Based on these data, we hypothesized that Belatacept would inhibit the development of DSA after lung transplantation, and that this would result in better freedom from CLAD and survival. We performed a pilot RCT to assess the feasibility of conducting a large scale RCT because experience with Belatacept in lung transplantation is very limited (15-17).

## METHODS

### Study Design and Medical Regimen

We performed a pilot 2-center open label phase II RCT to assess the feasibility of conducting a phase III multicenter RCT examining the efficacy and safety of Belatacept in lung transplantation. The study protocol is included in the Supplemental Material. The primary endpoint of this pilot study was the feasibility metric of randomizing 80% of eligible patients within 4 hours of completion of transplantation (Table 1). Secondary endpoints are listed in Table 1. We also sought to estimate the effect size of Belatacept on different clinical endpoints in this pilot study to inform the design and power calculations of a future pivotal clinical trial. Because cultures from bronchoscopy specimens are often positive for bacterial or fungal organisms, we defined infection as any positive culture result that required treatment. Our sites’ clinical protocols are to use antimicrobial therapy for any positive culture result from bronchoscopy specimens. However, we did not consider bronchoscopy cultures positive for *Candida* species as infections unless there was clinical evidence of invasive disease. We enrolled patients after listing for transplantation and randomized eligible participants after transplantation using a computer-generated block randomization method with a 1:1 ratio. Eligibility criteria for enrollment and randomization are listed in Table 2 and Table 3, respectively. Participants were randomized to either the Belatacept arm or the control arm. In the Belatacept arm, participants were treated with Belatacept, Tacrolimus, and prednisone starting on day 0 through day 89. On day 90, Tacrolimus was replaced by Mycophenolate Mofetil (MMF), and Belatacept and prednisone were continued through day 365. In the control arm, participants were treated with Tacrolimus, MMF, and prednisone from day 0 through day 365. In both study arms, Tacrolimus was initiated enterally or sublingually within the first 48 hours after transplantation and dosed to target a trough blood level of 8-15 ng/mL if kidney function was normal or 4-8 ng/mL if kidney function was impaired. MMF was dosed at 1 g twice daily. The first dose of Belatacept was given after transplant on day 0 at 10 mg/kg and subsequent doses were given at 10 mg/kg on days 7, 14, 28, 56, 84, then at 5 mg/kg on days 112, 140, 168, 196, 224, 252, 280, 308, 336 and 364. After the first 4 doses of Belatacept, subsequent doses were given within a 3-day window of the specified timepoints. All participants were treated with rabbit anti-thymocyte globulin (ATG) 1 mg/kg on days 0, 1, and 2 for induction immunosuppression. The first dose of Belatacept was given 12 hours after the first dose of ATG in participants randomized to Belatacept to avoid thrombotic complications. All participants were treated with methylprednisolone 500 mg intravenously before perfusion of the allograft during the transplant surgery. After transplant, participants were treated with methylprednisolone 0.5 mg/kg intravenously twice daily for 6 doses, then prednisone 0.5 mg/kg orally daily through day 14, then 0.2 mg/kg daily through day 30, then 0.1 mg/kg daily through day 180, then 5 mg daily through day 365. CMV seronegative recipients of seropositive donors or CMV seropositive recipients were treated with valganciclovir for CMV prophylaxis through day 365. CMV seronegative recipients of seronegative donors were treated with acyclovir for herpes and varicella prophylaxis. All participants received prophylaxis against *Pneumocystis jirovecii*. Antifungal prophylaxis was tailored according to culture results from bronchoscopy specimens.

**Table 1.**
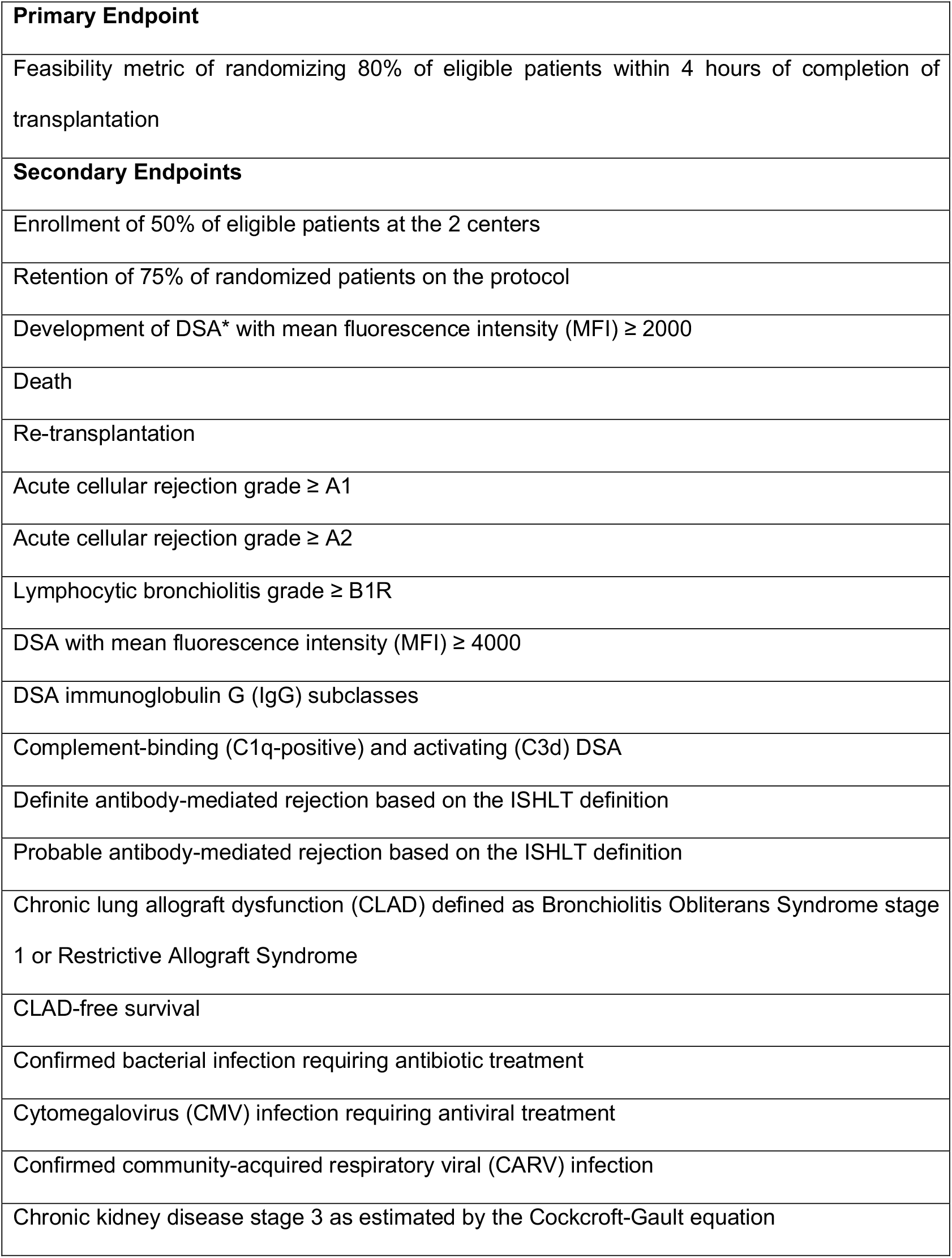
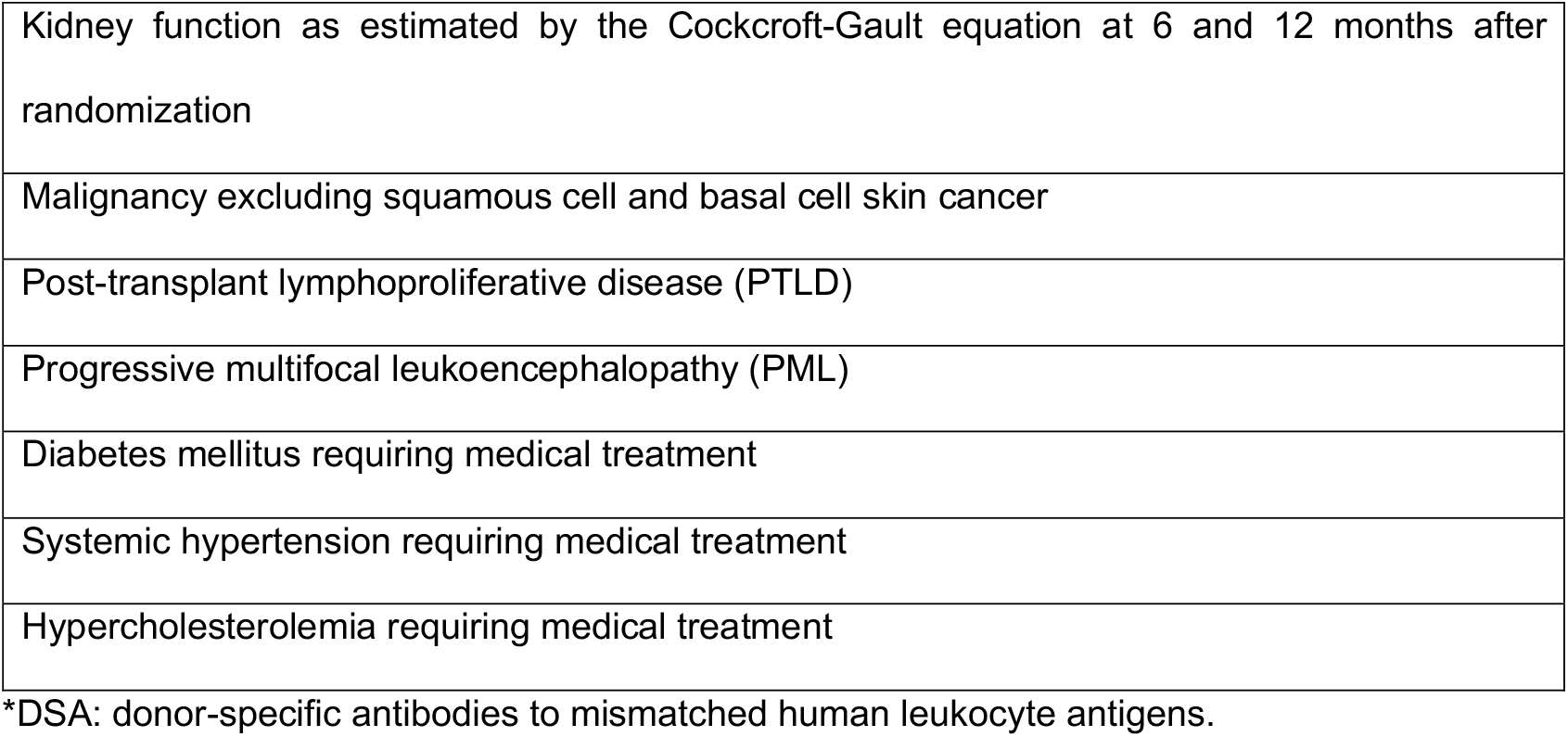
Study endpoints.

|  |
| --- |
| **Primary Endpoint** |
| Feasibility metric of randomizing 80% of eligible patients within 4 hours of completion of transplantation |
| **Secondary Endpoints** |
| Enrollment of 50% of eligible patients at the 2 centers |
| Retention of 75% of randomized patients on the protocol |
| Development of DSA* with mean fluorescence intensity (MFI) ≥ 2000 |
| Death |
| Re-transplantation |
| Acute cellular rejection grade ≥ A1 |
| Acute cellular rejection grade ≥ A2 |
| Lymphocytic bronchiolitis grade ≥ B1R |
| DSA with mean fluorescence intensity (MFI) ≥ 4000 |
| DSA immunoglobulin G (IgG) subclasses |
| Complement-binding (C1q-positive) and activating (C3d) DSA |
| Definite antibody-mediated rejection based on the ISHLT definition |
| Probable antibody-mediated rejection based on the ISHLT definition |
| Chronic lung allograft dysfunction (CLAD) defined as Bronchiolitis Obliterans Syndrome stage 1 or Restrictive Allograft Syndrome |
| CLAD-free survival |
| Confirmed bacterial infection requiring antibiotic treatment |
| Cytomegalovirus (CMV) infection requiring antiviral treatment |
| Confirmed community-acquired respiratory viral (CARV) infection |
| Chronic kidney disease stage 3 as estimated by the Cockcroft-Gault equation |
| Kidney function as estimated by the Cockcroft-Gault equation at 6 and 12 months after randomization |
| Malignancy excluding squamous cell and basal cell skin cancer |
| Post-transplant lymphoproliferative disease (PTLD) |
| Progressive multifocal leukoencephalopathy (PML) |
| Diabetes mellitus requiring medical treatment |
| Systemic hypertension requiring medical treatment |
| Hypercholesterolemia requiring medical treatment |

\*DSA: donor-specific antibodies to mismatched human leukocyte antigens.

**Table 2.**
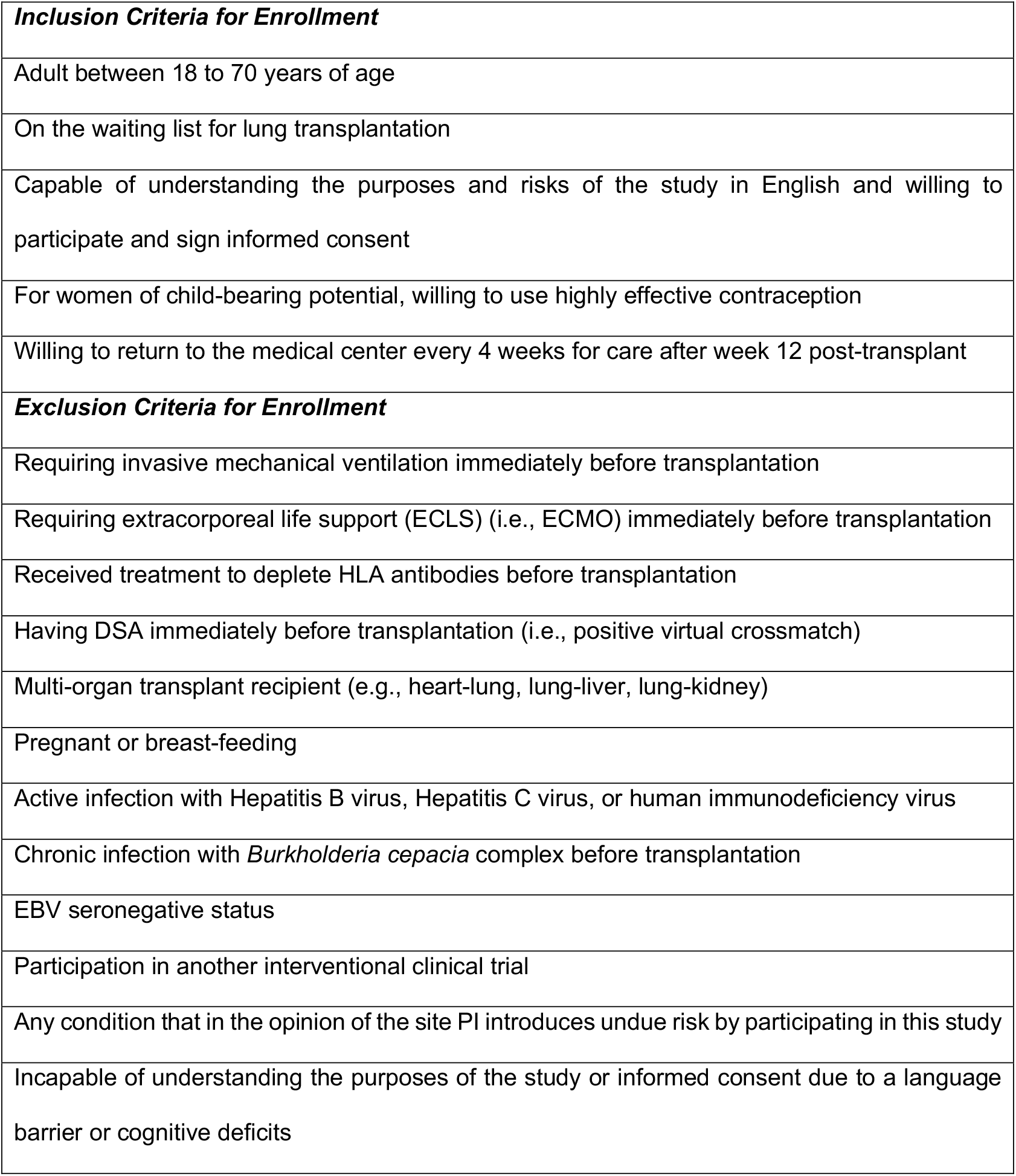
Eligibility criteria for enrollment.

**Table 3.**
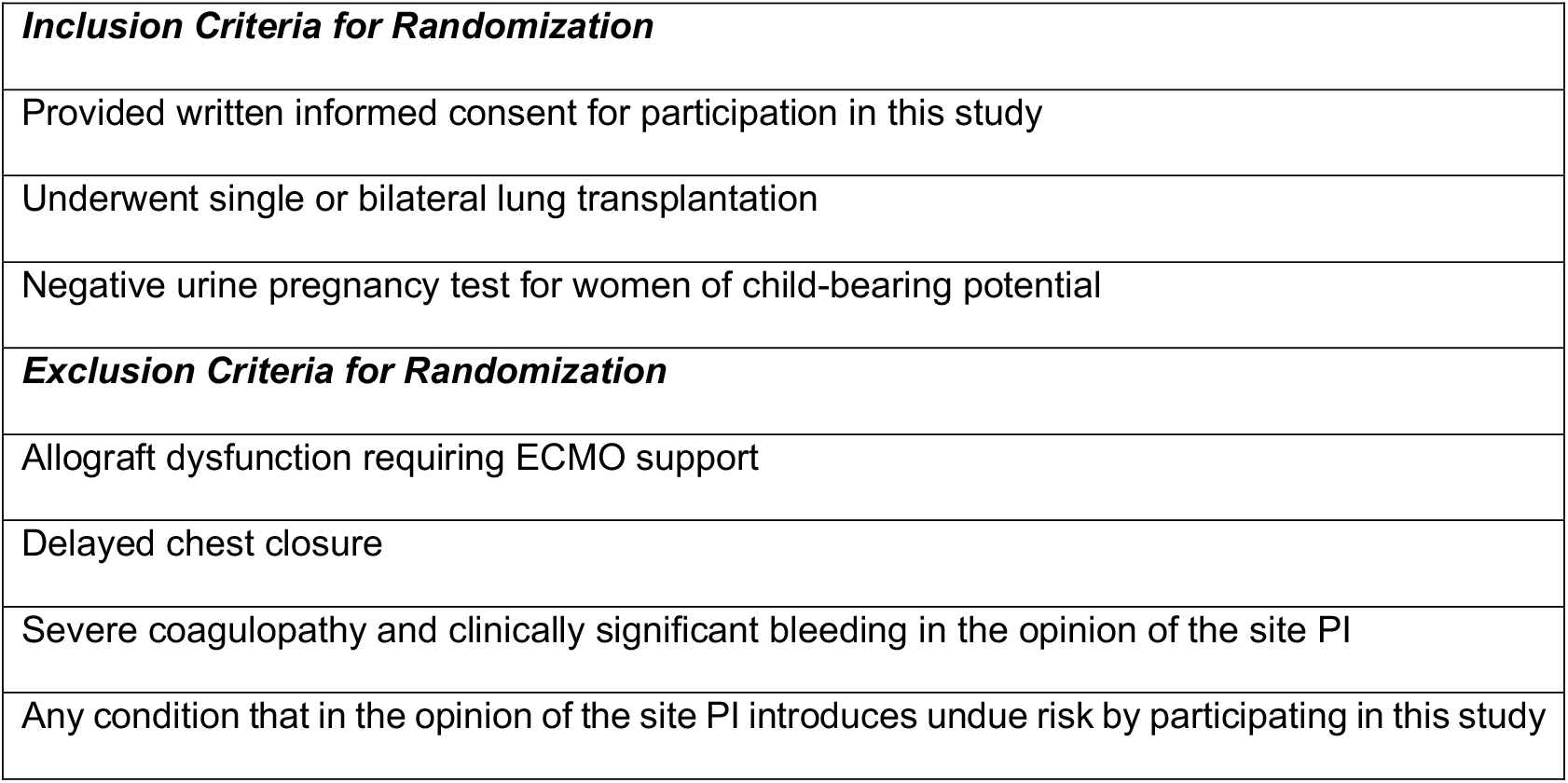
Eligibility criteria for randomization.

|  |
| --- |
| ***Inclusion Criteria for Randomization*** |
| Provided written informed consent for participation in this study |
| Underwent single or bilateral lung transplantation |
| Negative urine pregnancy test for women of child-bearing potential |
| ***Exclusion Criteria for Randomization*** |
| Allograft dysfunction requiring ECMO support |
| Delayed chest closure |
| Severe coagulopathy and clinically significant bleeding in the opinion of the site PI |
| Any condition that in the opinion of the site PI introduces undue risk by participating in this study |

### Testing

All participants underwent HLA typing using next generation sequencing (NGS) or reverse sequence specific oligonucleotide probes (rSSOP) based typing of HLA Loci A, B, C, DRB1, DRB3, DRB4, DRB5, DQA1, DQB1, DPA1 and DPB1. Participants were tested for HLA antibodies using the single antigen bead assay (One Lambda, a Thermo Fisher Company, Canoga Park, CA) before listing for transplantation. Donors were accepted only if the virtual crossmatch was negative, and a direct flow cytometry crossmatch was performed at the time of transplantation. Donors also underwent HLA typing at the sites after transplantation using NGS. Participants were followed at the sites’ clinics every 1-2 weeks in the first 3 months after transplantation then monthly for the duration of the study. These visits included a history and physical exam, routine labs, and pulmonary function tests. Bronchoscopy with bronchoalveolar lavage (BAL) and transbronchial lung biopsies was performed on days 28, 84, 112, 168, 252, and 365 (± 14 days) and if subjects developed signs or symptoms of allograft dysfunction. The study used a core HLA lab at Baylor University Medical Center for all post-transplant HLA antibody tests, and the study HLA investigator and technicians were blinded to participants’ study arm assignment. Samples were tested using the single antigen bead assay, and DSA was defined as reactivity with a mean fluorescence intensity (MFI) ≥ 2,000. Participants were tested for DSA on days 0, 10 (± 3 days), 28, 56, 84, 112, 168, 252, and 365 (± 14 days) and if they developed signs or symptoms of allograft dysfunction.

### Safety

The study protocol defined SAE as any untoward medical event that: results in death, is life-threatening, requires inpatient hospitalization or prolongs an existing hospitalization, results in persistent or significant disability or incapacity, results in birth defect, results in drug-induced liver injury, results in transmission of an infectious agent via the study drug, is an important medical event, pregnancy, cancer, or overdose. SAE were reported to Bristol Myers Squibb in real time, and any event that resulted in death was reported to the Data Safety Monitoring Board (DSMB), the US Food and Drug Administration (FDA), and the National Heart, Lung, and Blood Institute (NHLBI) safety officer within 1 day of the investigators’ receipt of this information. SAE, non-serious adverse events, protocol deviations, and data collection were reported to the DSMB during regularly scheduled meetings.

### Study Oversight

Funding for the study was provided through a grant from NHLBI (HL138186). Belatacept was provided at no cost by Bristol Myers Squibb through an Investigator Sponsored Research program (IM103-387). The study protocol was approved by the FDA, and Belatacept was used under an Investigational New Drug authorization (IND#138662). The sites’ local Institutional Review Boards (IRB) approved the study protocol. An independent DSMB approved the study protocol and monitored enrollment, data collection, and safety. The study was registered on ClinicalTrials.gov (NCT03388008). Protocol-specified individual participant stopping rules are listed in Table 4. Study-wide stopping rules are listed in Table 5. We assessed the rate of development of the study-wide stopping rule outcomes (Table 5) after every 10 participants were randomized and had at least 6 months of follow-up. If the incidence of any of these outcomes in the Belatacept arm was > 50%, and this was more than 50% higher than the incidence in the control arm, the protocol mandated that the investigators seek guidance from the DSMB about study continuation, suspension, or termination.

**Table 4.** Individual participant stopping rules.

|  |
| --- |
| Three episodes of acute cellular rejection grade ≥ A2 |
| Three episodes of lymphocytic bronchiolitis grade ≥ B1R |
| Development of chronic lung allograft dysfunction |
| Development of antibody-mediated rejection |
| Serious infection that in the opinion of the investigator warrants changing the maintenance immunosuppressive regimen |
| Malignancy including post-transplant lymphoproliferative disease, but excluding non-melanoma skin cancer |

**Table 5.**
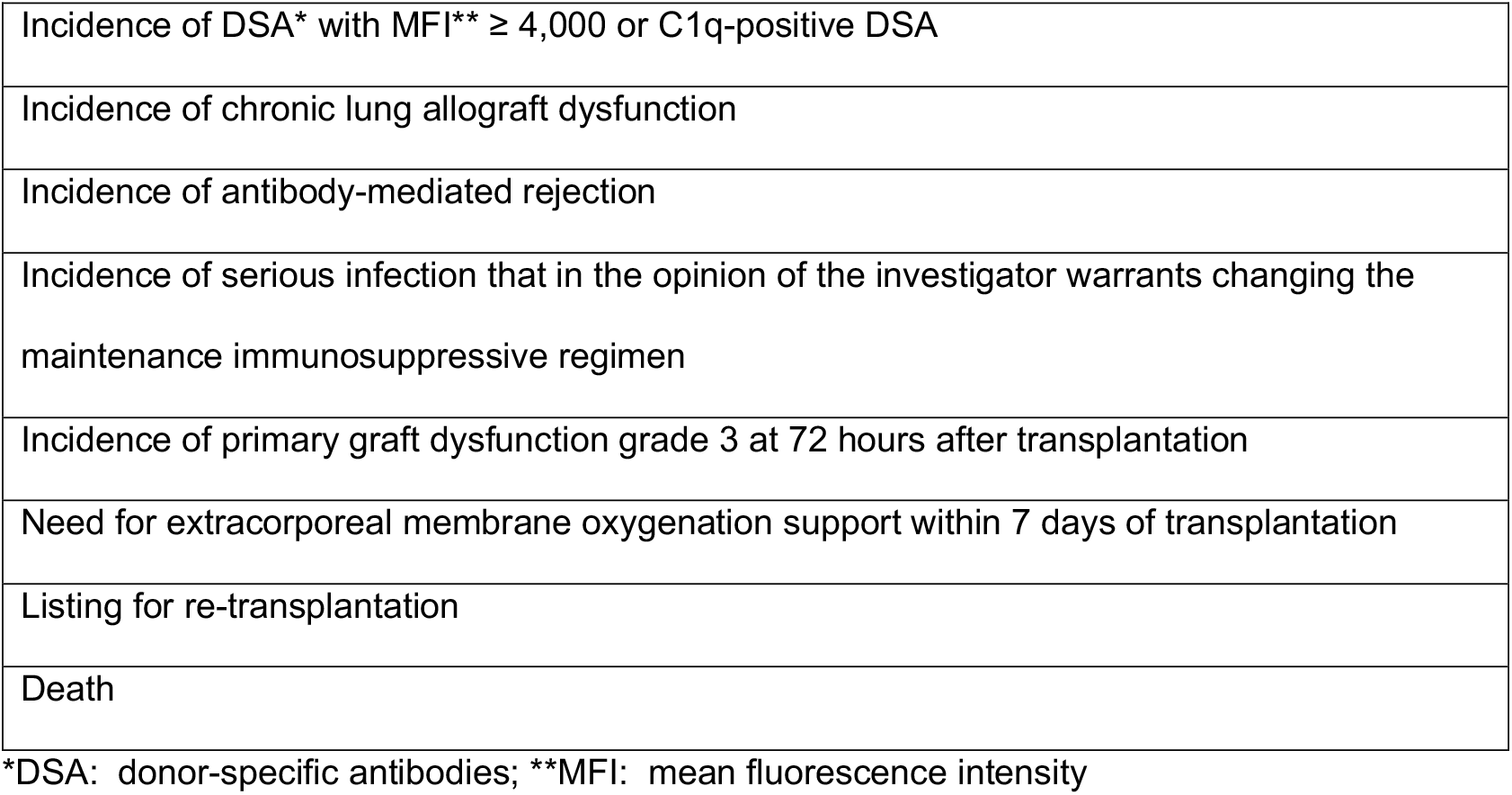
Study-wide stopping rules.

### Statistical Analysis

This pilot study was designed to assess the feasibility of conducting a large-scale phase III RCT. We planned to assess the randomization rate to inform the design of a future trial and aimed to randomize 40 subjects as this would generate 95% confidence intervals of ± 12% for the randomization rate. We did not expect that this pilot study would detect a statistically significant difference in clinical outcomes between the 2 groups because of the small sample size. We compared baseline characteristics between the 2 groups using t-tests (or Wilcoxon-Rank sum tests if the data were not normally distributed) and chi-square tests. We used the Kaplan-Meier method to report freedom from DSA, ACR, and survival and compared the outcomes between the 2 groups using the log rank test. We conducted the statistical analyses using SPSS and Prism and considered p < 0.05 statistically significant. We conducted all analyses according to the intention to treat principle.

## RESULTS

We began enrollment on December 1, 2019, and the first participant was randomized on January 2, 2020. As of May 30, 2021, 49 participants were enrolled, and 27 were randomized: 13 were randomized to Belatacept and 14 were randomized to Control (Figure 1). All subjects who were eligible for randomization were randomized within 4 hours of completion of transplantation. Baseline characteristics of the randomized participants are shown in Table 6. Between January 2, 2020, and May 30, 2021, 3 participants randomized to Belatacept died compared to none of the participants randomized to Control. After the 3^rd^ death occurred on May 30, 2021, we halted enrollment and randomization and notified the DSMB and FDA. The DSMB convened a meeting on June 3, 2021, to discuss study status. During the meeting, we proposed permanently terminating enrollment, randomization, and treatment because the risk-benefit ratio of continuing the study was no longer favorable. We also proposed ongoing follow-up of study participants for safety and efficacy reasons. The DSMB agreed that enrollment, randomization, and treatments should be permanently terminated, and ongoing follow-up was appropriate. Participants who were in the Belatacept arm were then converted to standard of care immunosuppression with Tacrolimus, MMF, and prednisone. Between June 3, 2021, and July 29, 2021, 2 additional subjects randomized to Belatacept died. As of August 30, 2021, 5 of the 13 subjects randomized to Belatacept died compared to none of the 14 randomized to Control (Figure 2, log rank p = 0.016). The causes of death are listed in Table 7. These include COVID-19 infection, restrictive allograft syndrome (RAS), hemothorax, suspected pulmonary embolism, and post-transplant lymphoproliferative disease. Of note, the subject who died of RAS developed rhinovirus/enterovirus respiratory infection with acute hypoxemic respiratory failure 6 weeks prior to the diagnosis of RAS. The subject who died of a hemothorax had received the third dose of Belatacept on the same day as the onset of the hemothorax (day 14 after transplant). The subject who died of a suspected pulmonary embolism had suffered a pelvic fracture after a fall and had been in a rehab facility before discharge home; he died suddenly at home.

**Figure 1.**
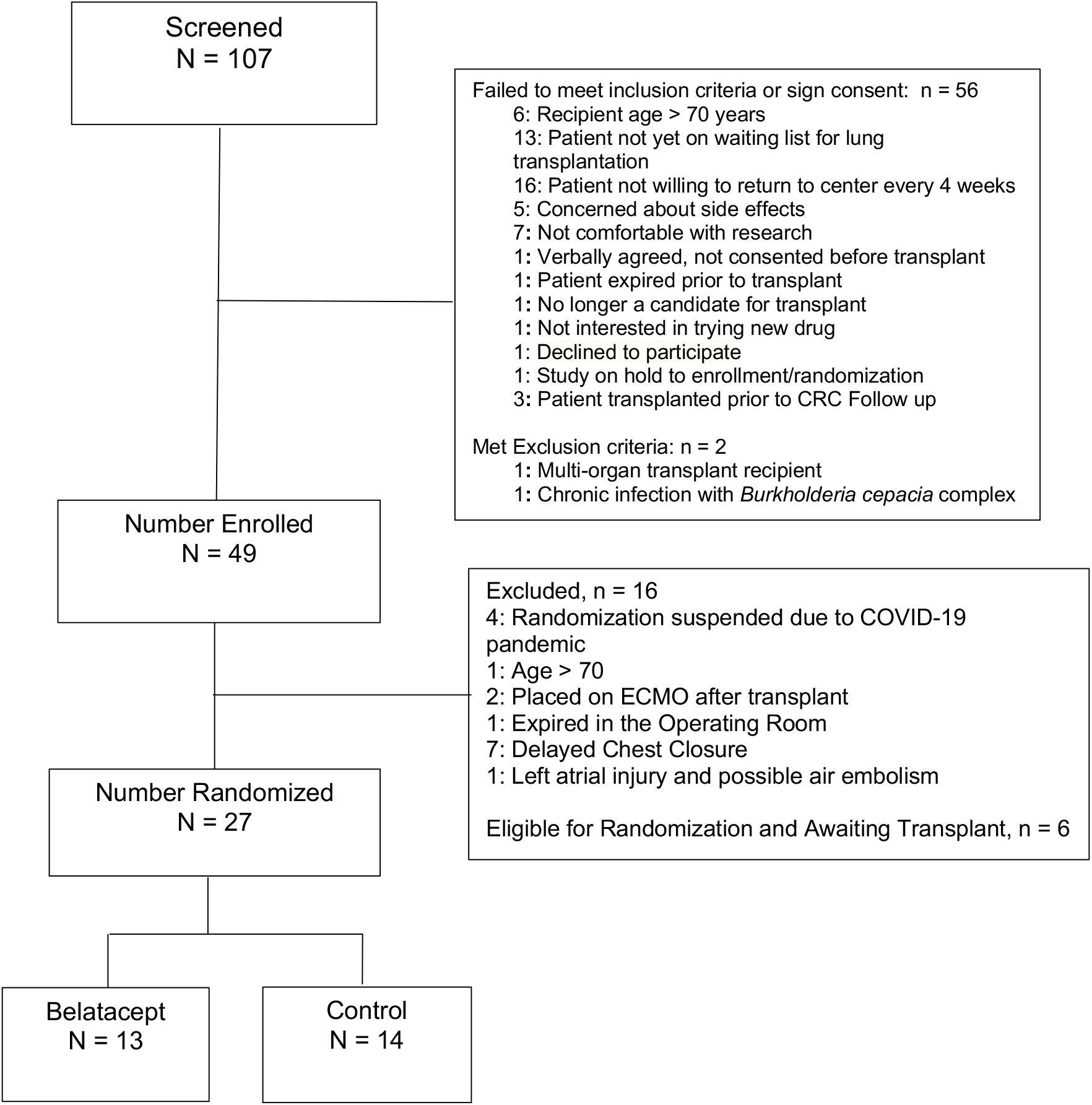
Study consort diagram.

**Figure 2.**
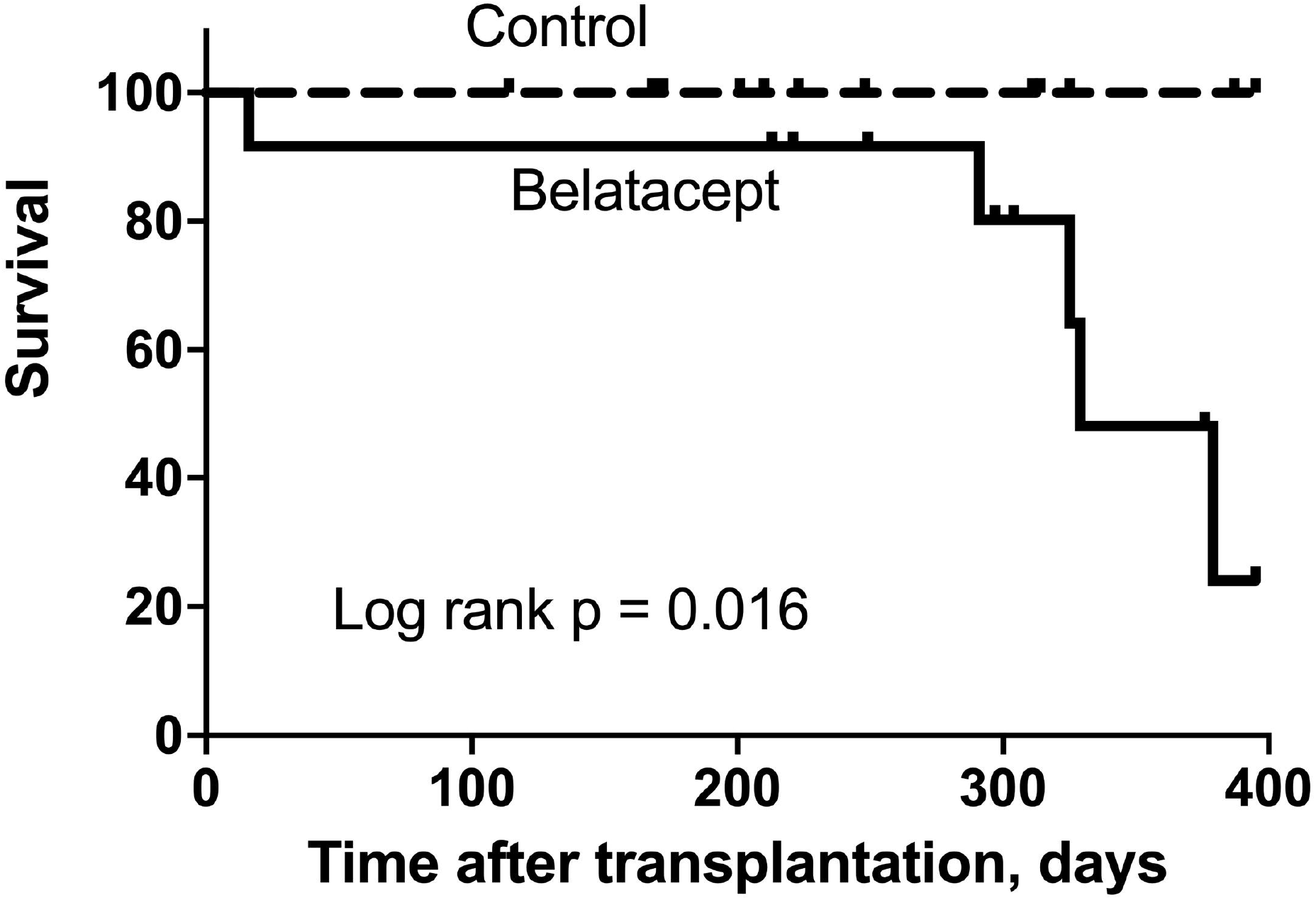
Patient survival. There was a significant survival difference between the Control group and the Belatacept group (log rank p = 0.016).

**Table 6.** Baseline characteristics of randomized participants.

|  | Belatacept<br>N = 13 | Control<br>N = 14 | p value |
| --- | --- | --- | --- |
| Recipient age, mean ± SD | 57.8 ± 7.5 | 59.5 ± 11.8 | 0.655 |
| Recipient gender |  |  | 0.785 |
| Female, n (%) | 4 (31%) | 5 (36%) |  |
| Male, n (%) | 9 (69%) | 9 (64%) |  |
| Recipient race |  |  | 0.586 |
| Black, n (%) | 1 (8%) | 2 (14%) |  |
| White, n (%) | 12 (92%) | 12 (86%) |  |
| Recipient ethnicity |  |  | 0.957 |
| Hispanic, n (%) | 1 (8%) | 1 (7%) |  |
| Not Hispanic, n (%) | 12 (92%) | 13 (93%) |  |
| Diagnosis leading to transplantation |  |  | 0.202 |
| Interstitial lung disease, n (%) | 6 (46%) | 8 (57%) |  |
| Chronic obstructive pulmonary disease, n (%) | 4 (31%) | 0 |  |
| Pulmonary arterial hypertension, n (%) | 0 | 2 (14%) |  |
| Bronchiectasis, n (%) | 1 (8%) | 1 (7%) |  |
| Sarcoidosis, n (%) | 0 | 1 (7%) |  |
| Other, n (%) | 2 (15%) | 2 (14%) |  |
| Operation |  |  | 0.315 |
| Single lung transplant, n (%) | 1 (8%) | 3 (21%) |  |
| Bilateral lung transplant, n (%) | 12 (92%) | 11 (79%) |  |

**Table 7.**
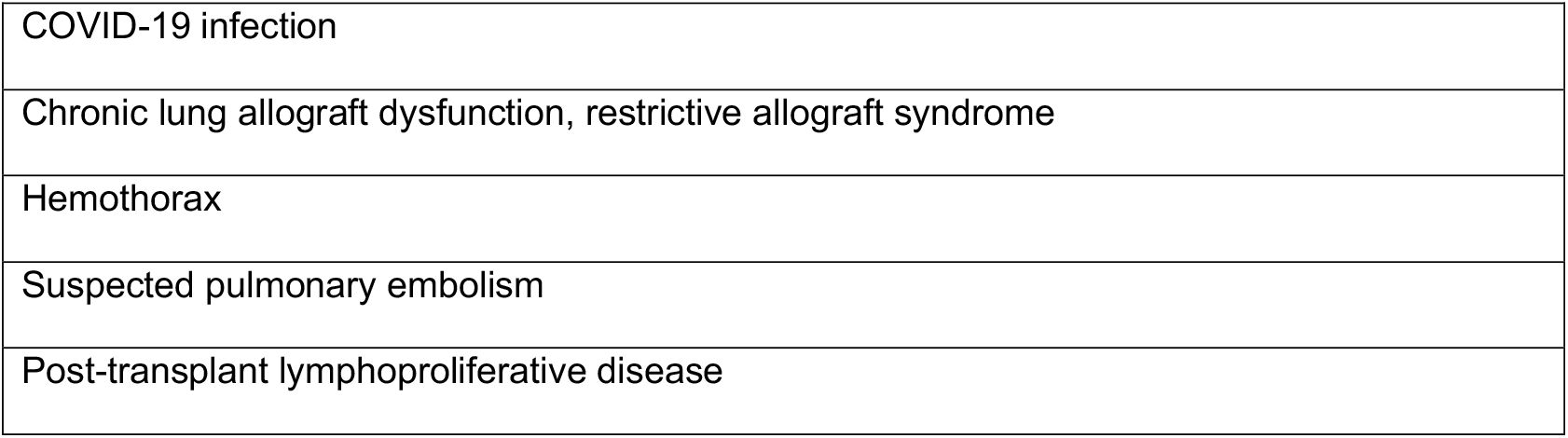
Causes of death.

|  |
| --- |
| COVID-19 infection |
| Chronic lung allograft dysfunction, restrictive allograft syndrome |
| Hemothorax |
| Suspected pulmonary embolism |
| Post-transplant lymphoproliferative disease |

Using an MFI threshold ≥ 2,000 to identify DSA, 6 of the 13 participants randomized to Belatacept and 7 of the 14 randomized to Control developed DSA (p= 0.842), and there was no difference in freedom from DSA between the 2 groups (Figure 3A, log rank p = 0.908). There was also no significant difference in the DSA class between the 2 groups (p = 0.131): among those in the Belatacept arm, 3 developed DSA only to class I HLA, 1 developed DSA only to class II HLA, 2 developed DSA to class I and II HLA, and 3 developed DSA to HLA-DQ; among those in the Control arm, 5 developed DSA only to class II HLA, 2 developed DSA to class I and II HLA, and 5 developed DSA to HLA-DQ (Figure 3B). Using an MFI threshold ≥ 4,000 to identify DSA, 3 of the 13 participants randomized to Belatacept and 4 of the 14 randomized to Control developed DSA (p = 0.745), and there was no difference in freedom from DSA between the 2 groups (Figure 3C, log rank = 0.803). There was also no significant difference in the DSA class between the 2 groups (p = 0.745): among those in the Belatacept arm, 3 developed DSA only to class II HLA, and 3 developed DSA to HLA-DQ; among those in the Control arm, 4 developed DSA only to class II HLA, and 3 developed DSA to HLA-DQ (Figure 3D). Finally, 3 participants randomized to Belatacept and 4 randomized to Control developed C1q-positive DSA (p = 0.745).

**Figure 3.**
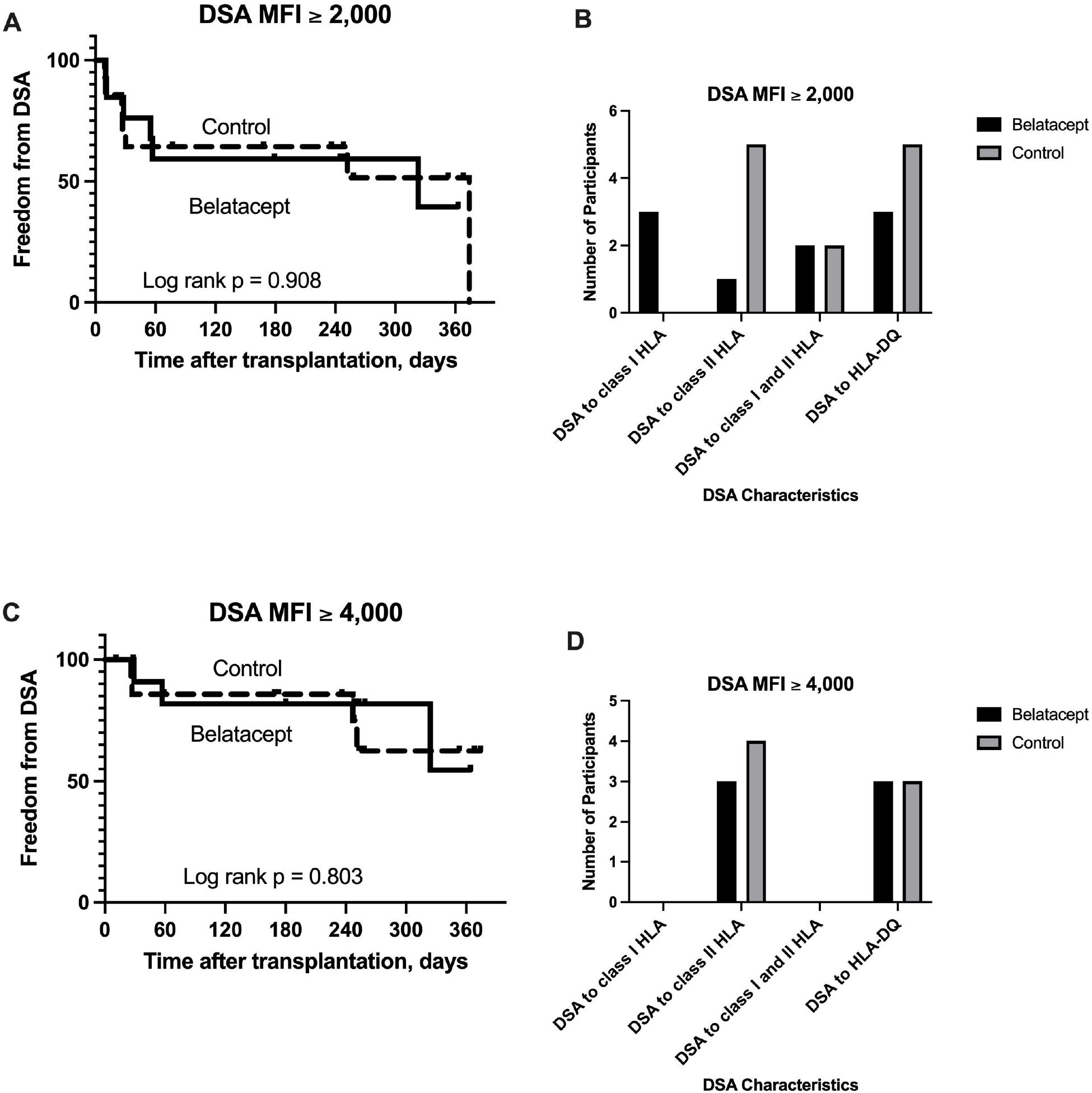
The development of donor-specific antibodies (DSA) to mismatched human leukocyte antigens (HLA). There was no significant difference in freedom from the development of DSA between the 2 groups.

During the follow-up period, there was no significant difference in the number of transbronchial lung biopsies performed in each group (p = 0.676). Those in the Belatacept arm underwent a median 4 transbronchial lung biopsies (mean ± SD = 3.9 ± 2.3), and those in the Control arm underwent a median 4 transbronchial lung biopsies (mean ± SD = 4.3 ± 2.1). There were no episodes of ACR grade A2 or higher during the follow-up period in either study arm. Four participants randomized to Belatacept developed at least 1 episode of ACR grade A1 compared to 5 participants randomized to Control (p = 0.785), and there was no significant difference in freedom from ACR grade A1 between the 2 groups (Figure 4A, log rank p = 0.702). Two participants in each group had more than 1 episode of ACR grade A1 during the follow-up period (p = 0.936). One participant in the Belatacept arm and 2 in the Control arm developed lymphocytic bronchiolitis grade B1R (p = 0.586), and there was no significant difference in freedom from lymphocytic bronchiolitis between the 2 groups (Figure 4B, log rank p = 0.634).

**Figure 4.**
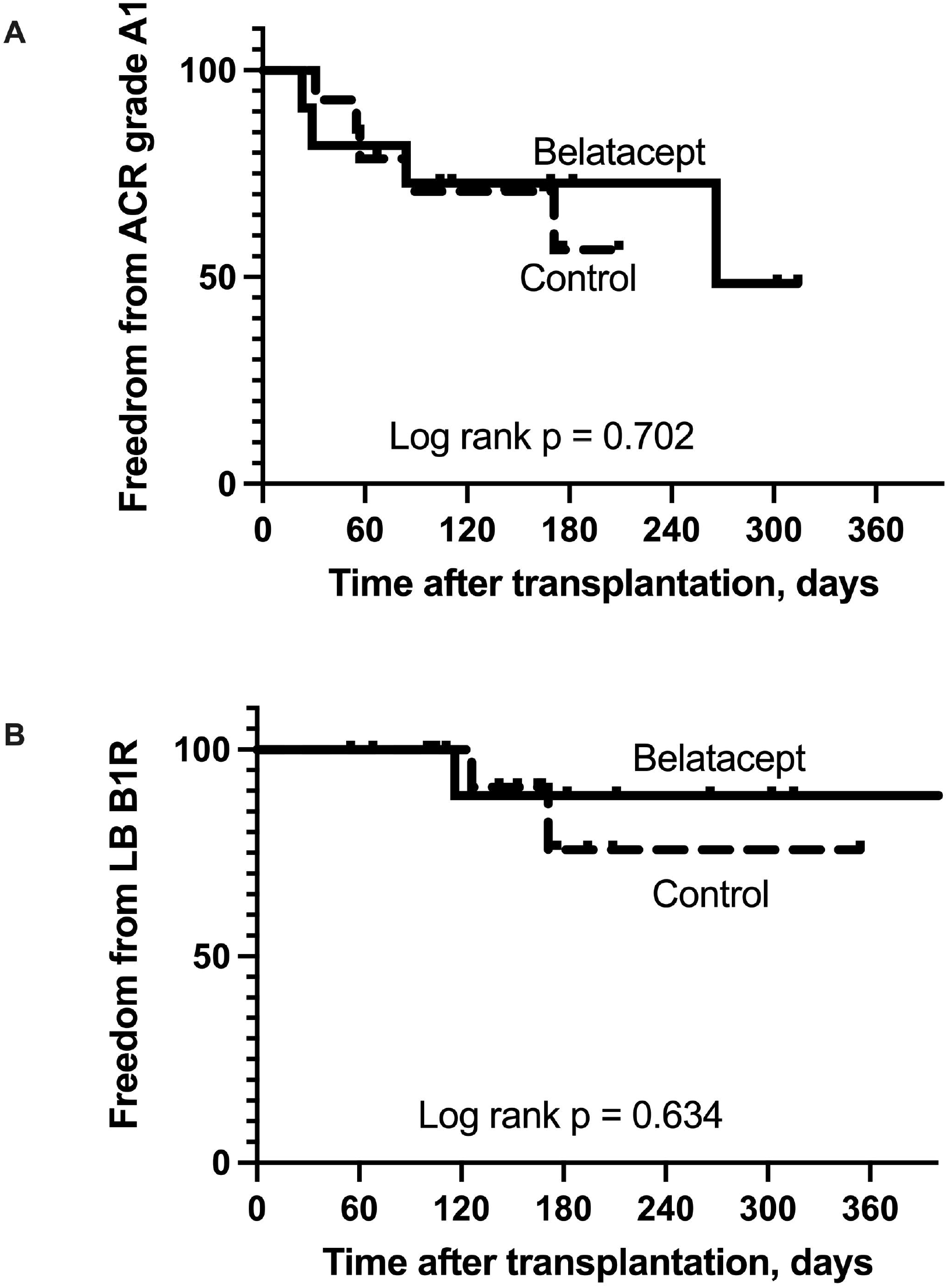
Acute cellular rejection and lymphocytic bronchiolitis. There was no significant difference in freedom from acute cellular rejection or lymphocytic bronchiolitis between the 2 groups.

Eleven participants randomized to Belatacept and 9 randomized to Control had at least 1 infection during the follow-up period (Table 8, p = 0.228). In total, there were 24 infections in the Belatacept group and 26 in the Control group. Bacterial respiratory tract infections were most common; 5 participants in the Belatacept arm had 8 bacterial infections, and 6 participants in the Control arm had 9 bacterial infections (Table 8, p = 0.816). Fungal respiratory tract infections were less common; 3 participants in the Belatacept arm had 4 fungal infections, and 6 in the Control arm had 6 fungal infections (p = 0.276). None of these bacterial or fungal infections was life-threatening or fatal. Two participants randomized to Belatacept and 3 randomized to Control developed COVID-19 infection (p = 0.686). This was fatal in 1 participant randomized to Belatacept whereas the remaining participants recovered without apparent sequelae.

**Table 8.**
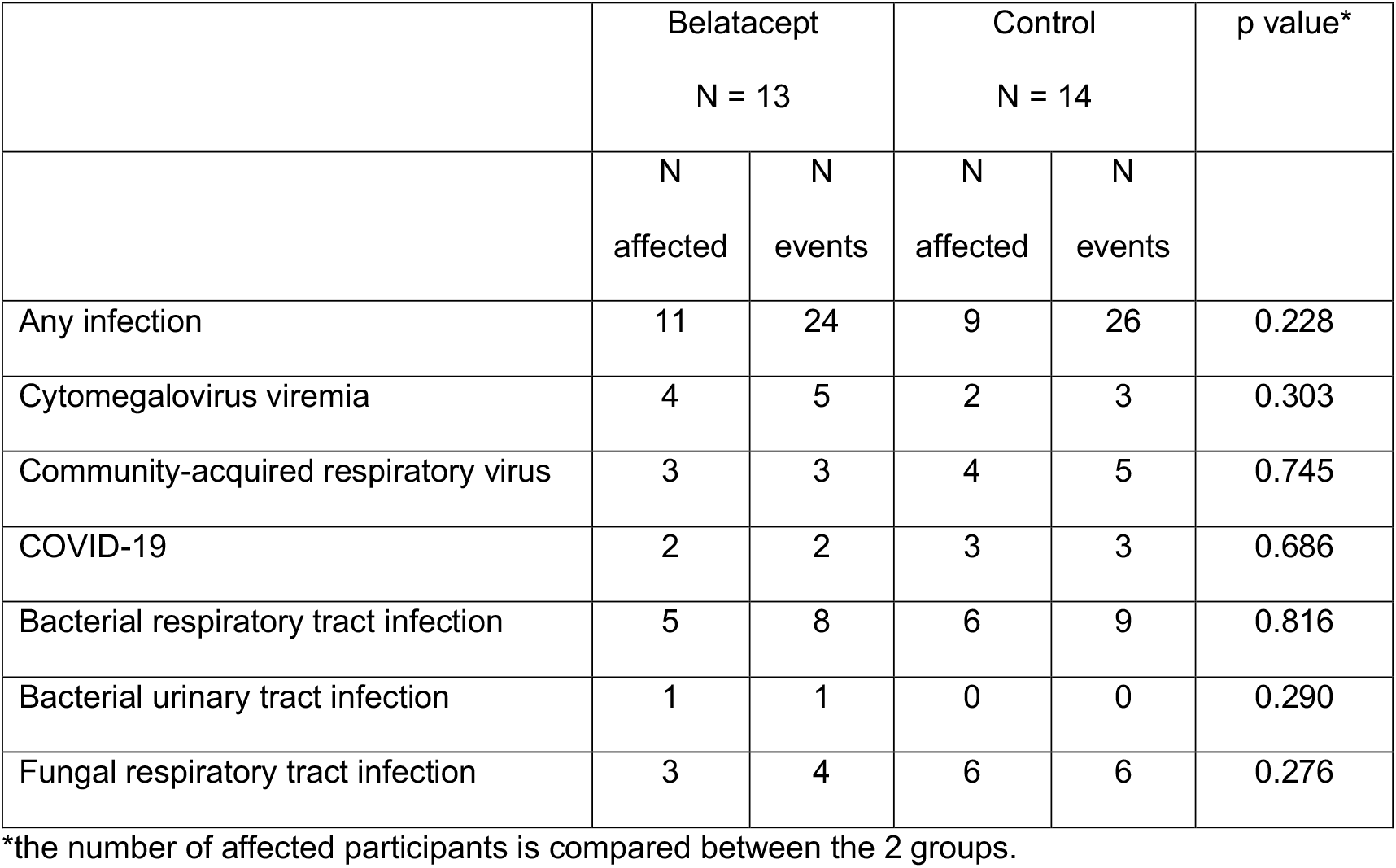
Infections.

|  | Belatacept<br>N = 13 |  | Control<br>N = 14 |  | p value* |
| --- | --- | --- | --- | --- | --- |
|  | N<br>affected | N<br>events | N<br>affected | N<br>events |  |
| Any infection | 11 | 24 | 9 | 26 | 0.228 |
| Cytomegalovirus viremia | 4 | 5 | 2 | 3 | 0.303 |
| Community-acquired respiratory virus | 3 | 3 | 4 | 5 | 0.745 |
| COVID-19 | 2 | 2 | 3 | 3 | 0.686 |
| Bacterial respiratory tract infection | 5 | 8 | 6 | 9 | 0.816 |
| Bacterial urinary tract infection | 1 | 1 | 0 | 0 | 0.290 |
| Fungal respiratory tract infection | 3 | 4 | 6 | 6 | 0.276 |

\*the number of affected participants is compared between the 2 groups.

During the follow-up period, 10 participants randomized to Belatacept and 10 randomized to Control experienced at least 1 SAE (p = 0.745). The most common SAE was acute hypoxemic respiratory failure. Six participants randomized to Belatacept experienced 12 episodes of acute hypoxemic respiratory failure; 1 participant experienced 6 episodes of respiratory failure. Four participants randomized to Control experienced 4 episodes of respiratory failure (Table 9). Common causes of respiratory failure in both arms included respiratory infection, early post-operative respiratory failure, and airway complications. Other SAE are listed in Table 9; only events that met the definition of SAE are listed here.

**Table 9.**
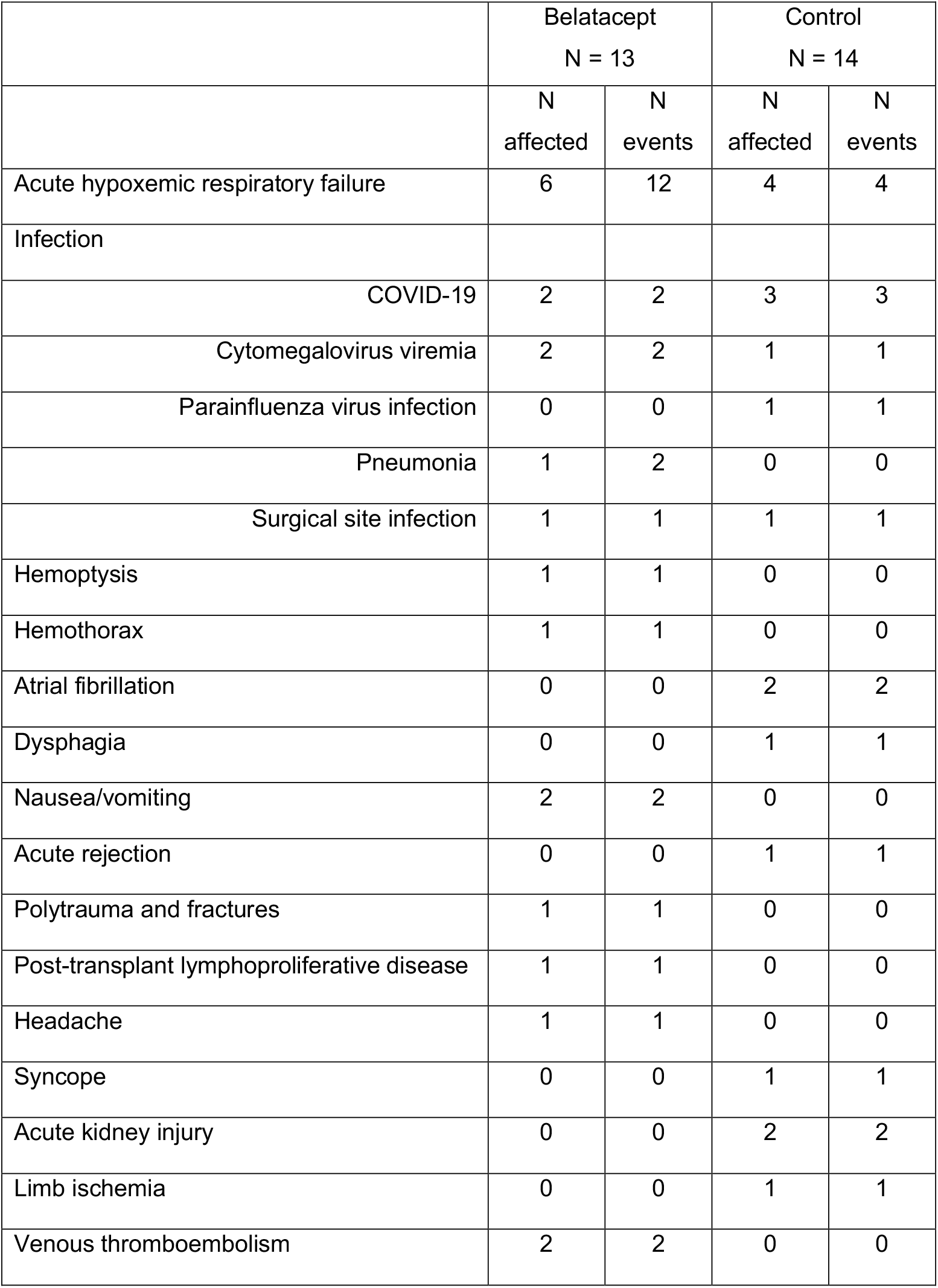
Serious adverse events.

|  | Belatacept<br>N = 13 |  | Control<br>N = 14 |  |
| --- | --- | --- | --- | --- |
|  | N<br>affected | N<br>events | N<br>affected | N<br>events |
| Acute hypoxemic respiratory failure | 6 | 12 | 4 | 4 |
| Infection |  |  |  |  |
| COVID-19 | 2 | 2 | 3 | 3 |
| Cytomegalovirus viremia | 2 | 2 | 1 | 1 |
| Parainfluenza virus infection | 0 | 0 | 1 | 1 |
| Pneumonia | 1 | 2 | 0 | 0 |
| Surgical site infection | 1 | 1 | 1 | 1 |
| Hemoptysis | 1 | 1 | 0 | 0 |
| Hemothorax | 1 | 1 | 0 | 0 |
| Atrial fibrillation | 0 | 0 | 2 | 2 |
| Dysphagia | 0 | 0 | 1 | 1 |
| Nausea/vomiting | 2 | 2 | 0 | 0 |
| Acute rejection | 0 | 0 | 1 | 1 |
| Polytrauma and fractures | 1 | 1 | 0 | 0 |
| Post-transplant lymphoproliferative disease | 1 | 1 | 0 | 0 |
| Headache | 1 | 1 | 0 | 0 |
| Syncope | 0 | 0 | 1 | 1 |
| Acute kidney injury | 0 | 0 | 2 | 2 |
| Limb ischemia | 0 | 0 | 1 | 1 |
| Venous thromboembolism | 2 | 2 | 0 | 0 |

## DISCUSSION

This pilot RCT is the first to examine the role of Belatacept in lung transplantation. The results demonstrate a significantly higher mortality among participants randomized to Belatacept compared to those randomized to Control. Although it is difficult to understand the causes of this increased mortality, the regimen we used in this trial consisting of ATG induction and the Belatacept dosing schedule is clearly associated with an increased risk of death. Indeed, mortality among those in the Belatacept arm is considerably higher than the sites’ 1-year mortality rate outside this clinical trial and that reported by the ISHLT Registry, emphasizing that this increased mortality is due to the investigational regimen (1, 18, 19). In designing this trial, we were concerned about the risk of early ACR that was seen in kidney transplantation especially since ACR is more common after lung transplantation. Therefore, we included ATG induction to ameliorate the risk of ACR, and in fact, we did not detect an increased risk of ACR in this study. Obviously, we are unable to discern whether this was because of ATG induction or the maintenance immunosuppressive regimen. In addition, we did not identify a difference in the development of DSA or infections between the 2 groups. The lack of a clear association between Belatacept and ACR, DSA, or infection makes it challenging to determine whether this regimen resulted in “over-immunosuppression” or “under-immunosuppression.” Indeed, such a conclusion may be overly simplistic as lung transplant recipients are inherently complex with diverse pre- and post-transplant co-morbidities that may impact outcomes.

Single-center retrospective studies of Belatacept in lung transplantation have generally supported its use as a rescue treatment for calcineurin inhibitor toxicity although there is one case report of fulminant allograft failure after conversion to Belatacept (15-17). There are numerous differences between these studies and ours including study design and eligibility criteria. We used Belatacept as *de novo* immunosuppression immediately after transplantation and used ATG induction for all participants in this study. In contrast, many patients included in previous studies had undergone transplantation months or years earlier, and the risk of serious complications of immunosuppression is likely related to the timepoint after transplantation. Nonetheless, it is noteworthy that patients included in previous reports are inherently at increased risk of complications because of the toxicities that led to the initiation of Belatacept. Our results are similar to those reported by a multicenter RCT in liver transplantation which demonstrated increased mortality and allograft loss in 2 of 3 Belatacept groups compared to standard of care (20). In this RCT of Belatacept in liver transplantation, all Belatacept groups had higher rates of ACR and viral and fungal infections (20). Although we did not observe an increased risk of ACR or infection in our study, it is possible that this is related to the small sample size as our pilot RCT was not powered to detect differences in clinical outcomes. It is also possible that our data underestimate the true incidence of DSA, ACR, and infection in the Belatacept arm because of the excess mortality and shorter duration of follow-up among subjects who died.

We conclude that the investigational regimen used in this RCT consisting of this Belatacept dosing schedule and ATG induction results in increased mortality after lung transplantation. Clearly, we are unable to comment on the use of Belatacept without ATG induction or the safety and efficacy of a different Belatacept dosing schedule. Nonetheless, our results caution against the routine use of Belatacept in lung transplantation. These findings underscore the need to examine novel therapies in lung transplantation in the context of carefully designed and monitored randomized controlled trials.

## Supporting information

Supplemental Materials and Study Protocol

## Data Availability

All data produced in the present work are contained in the manuscript.

