## Supplemental Materials and Study Protocol for "A Pilot Randomized Controlled Trial of *de novo* Belatacept-Based Immunosuppression in Lung Transplantation"

#### SUPPLEMENTAL MATERIAL

#### SUMMARY OF PROTOCOL MODIFICATIONS

Protocol version 15.0 was approved by the DSMB and the sites' IRBs before enrollment began in December 2019. After the first DSMB meeting in October 2020, the protocol was modified to clarify that the investigators may discontinue Belatacept if a participant developed an AE, an SAE, or any medical condition that would place the participant at undue risk by continuing Belatacept, and that such participants would continue to be followed by the study for safety and efficacy endpoints (Protocol version 16.0). In February 2021, the DSA Study-Wide Stopping Rule was met when DSA was detected in 4 of 6 participants randomized to Belatacept and 0 of 4 participants randomized to Control when DSA was defined as any donor-specific reactivity with MFI ≥ 1,000. As a result, we sought guidance from the DSMB about study continuation, suspension, or termination. The DSMB recommended modifying the DSA Study-Wide Stopping Rule to a more clinically significant threshold. Consequently, we modified the DSA definition to donor-specific reactivity with MFI ≥ 2,000 and modified the Stopping Rule to the development of DSA with MFI ≥ 4,000 or C1q-positive DSA (Protocol version 17.0). Protocol version 17.0 is below.

### A PILOT RANDOMIZED CONTROLLED TRIAL OF DE NOVO BELATACEPT-BASED IMMUNOSUPPRESSION IN LUNG TRANSPLANTATION

Howard J. Huang, MD
Ramsey R. Hachem, MD

Study ID: IM103-387
IND Number: 138662
ClinicalTrials.gov Identifier: NCT03388008

Draft: v 17.0
Date: 1 March 2021

#### TABLE OF CONTENTS


| Section | Page |
|---|---|
| 1. List of Abbreviations | 3 |
| 2. Protocol Summary | 4 |
| 3. Study Schematic | 4 |
| 4. Key Roles | 5 |
| 5. Introduction | 6 |
| 6. Objectives and Purpose | 8 |
| 7. Study Design and Endpoints | 9 |
| 8. Study Enrollment | 10 |
| 9. Study Agent | 13 |
| 10. Study Procedures and Schedule | 14 |
| 11. Safety Considerations | 18 |
| 12. Statistical Considerations | 23 |
| 13. References | 24 |

#### LIST OF ABBREVIATIONS

| | |
|---|---|
| ACR | Acute Cellular Rejection |
| AE | Adverse Event |
| AMR | Antibody-Mediated Rejection |
| ATG | Anti-Thymocyte Globulin |
| ATS | American Thoracic Society |
| BAL | Bronchoalveolar Lavage |
| BELA | Belatacept |
| BMS | Bristol-Myers Squibb |
| BOS | Bronchiolitis Obliterans Syndrome |
| CARV | Community-Acquired Respiratory Virus/Viral |
| CBC | Complete Blood Counts |
| CFR | Code of Federal Regulations |
| CIP | Cellular Immunophenotyping |
| CKD | Chronic Kidney Disease |
| CLAD | Chronic Lung Allograft Dysfunction |
| CMP | Comprehensive Metabolic Panel |
| CMV | Cytomegalovirus |
| CNI | Calcineurin Inhibitor |
| DCC | Data Coordinating Center |
| DSA | Donor-Specific HLA Antibodies |
| DSMB | Data Safety and Monitoring Board |
| EBV | Epstein-Barr Virus |
| ECLS | Extra-Corporeal Life Support |
| ECMO | Extra-Corporeal Membrane Oxygenation |
| FOB | Fiberoptic Bronchoscopy |
| GCP | Good Clinical Practice |
| HIV | Human Immunodeficiency Virus |
| HLA | Human Leukocyte Antigens |
| HSV | Herpes Simplex Virus |
| IgG | Immunoglobulin G |
| IP | Investigational Product |
| IRB | Institutional Review Board |
| IS | Immunosuppression |
| ISHLT | International Society for Heart and Lung Transplantation |
| LB | Lymphocytic Bronchiolitis |
| MMF | Mycophenolate mofetil |
| MPA | Mycophenolic acid |
| PCR | Polymerase Chain Reaction |
| PGD | Primary Graft Dysfunction |
| PTLD | Post-Transplant Lymphoproliferative Disease |
| RAS | Restrictive Allograft Syndrome |
| RCT | Randomized Controlled Trial |
| SAB | Single Antigen Bead |
| SAE | Serious Adverse Event |
| TBBX | Transbronchial lung Biopsy |
| TMP/SMX | Trimethoprim/sulfamethoxazole |
| VZV | Varicella Zoster Virus |

#### PROTOCOL SUMMARY

| | |
|---|---|
| Title | A Pilot Randomized Controlled Trial Of De Novo Belatacept-Based Immunosuppression In Lung Transplantation |
| Précis | This open-label, pilot, randomized controlled trial (RCT) will compare the combination of Belatacept, mycophenolate mofetil (MMF) and prednisone to tacrolimus, MMF, and prednisone after lung transplantation. The primary endpoint is a composite of the development of donor-specific HLA antibodies (DSA), death, or re-transplantation. |
| Objectives | The primary objective of this pilot study is to assess the feasibility of conducting a phase III multicenter RCT examining the efficacy and safety of Belatacept-based immunosuppression after lung transplantation. |
| Endpoints | The primary endpoint of the study is the feasibility metric of randomizing 80% of patients eligible for randomization within 4 hours of completion of transplantation.<br>Secondary endpoints include enrollment of 50% of patients eligible for enrollment, retention of 75% of randomized patients on the protocol, the development of DSA, death, re-transplantation, antibody-mediated rejection, acute cellular rejection, chronic lung allograft dysfunction, renal function, and infection 1 year after transplantation. |
| Population | 40 adult lung transplant recipients |
| Phase | II |
| Number of Sites | 2 sites:<br>• Houston Methodist Hospital<br>• Washington University School of Medicine in St. Louis |
| Study Agent | Belatacept 10 mg/kg on days 0, 7, 14, 28, 56, 84, then 5 mg/kg on days 112, 140, 168, 196, 224, 252, 280, 308, 336, and 364 |
| Study Duration | 3 years |
| Participant Duration | 1 year + 30 day follow-up |

#### STUDY SCHEMATIC

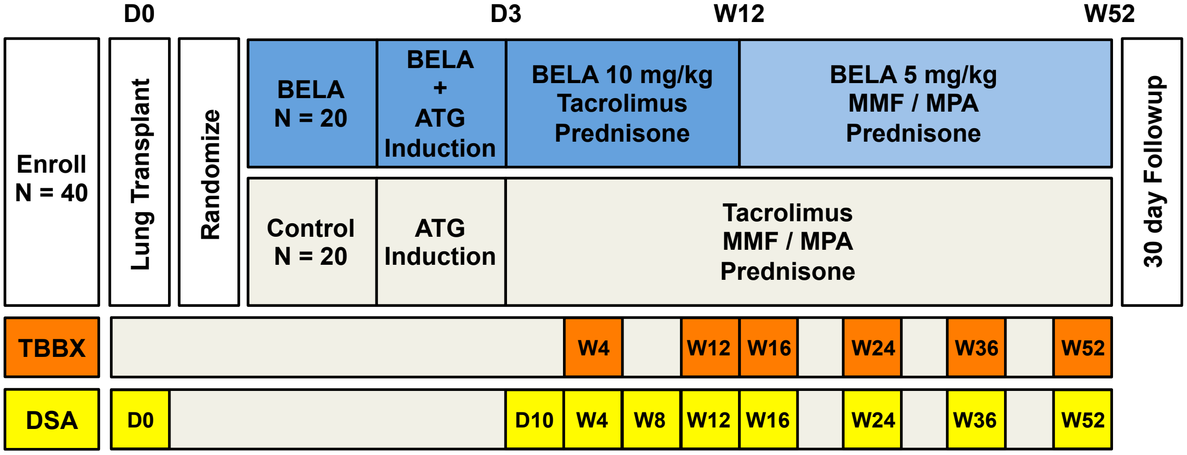

#### 1. KEY ROLES

**Co-Principal Investigator**
Howard J. Huang, MD
Chief, Section of Lung Transplantation
Houston Methodist J.C. Walter Jr. Transplant Center
6550 Fannin Street
Smith Tower, Suite 1001
Houston, TX 77030


**Co-Principal Investigator and Clinical Coordinating Center Director**
Ramsey R. Hachem, MD
Medical Director, Lung Transplantation
Professor of Medicine
Washington University School of Medicine in St. Louis
4523 Clayton Ave., Campus Box 8052
St. Louis, MO 63110


**Co-Principal Investigator and Data Coordinating Center Director**
Kenneth B. Schechtman, PhD
Associate Professor of Biostatistics and Medicine
Washington University School of Medicine
4523 Clayton Ave., Campus Box 8067
St. Louis, MO 63110-1093


**Histocompatibility Core Lab Investigator**
Medhat Askar, MD, PhD
Transplant Immunology Lab Director
Baylor University Medical Center at Dallas
3500 Gaston Ave. 4th Floor Y-Wing
Dallas, TX 75246


**Pathology Core Lab Investigator**
Joseph P. Gaut, MD, PhD
Department of Pathology and Immunology
Washington University School of Medicine
4523 Clayton Ave., Campus Box 8118
St. Louis, MO 63110


#### 2. INTRODUCTION

##### 2.1. BACKGROUND INFORMATION

Belatacept is a selective costimulation blocker that prevents T-cell activation. Belatacept is a fusion protein consisting of high-affinity receptor for CD80 and CD86 expressed on the surface of antigen-presenting cells and the Fc portion of human IgG. By binding CD80 and CD86, Belatacept blocks the second signal of T-cell activation thereby promoting anergy.

##### 2.2. RATIONALE

Lung transplantation is the ultimate treatment for patients with end-stage lung disease, and approximately 2000 patients undergo lung transplantation annually in the United States. However, long-term outcomes after transplantation remain disappointing, and the median survival after transplantation is approximately 5.5 years (1). Infection and primary graft dysfunction (PGD) are the most common causes of death in the first year after transplantation, but chronic lung allograft dysfunction (CLAD) is the leading cause of death beyond the first year (1). Different clinical phenotypes of CLAD have been defined including classic bronchiolitis obliterans syndrome (BOS) and restrictive allograft syndrome (RAS), and the incidence of CLAD approaches 50% within 3 years of transplantation (2-4). There is no effective evidence-based treatment for CLAD, and it typically follows a progressive clinical course ultimately resulting in allograft failure and death (4-6). Indeed, the median survival after the diagnosis of CLAD is approximately 3 years, and studies have consistently shown that patients who develop CLAD have a significantly worse quality of life (6, 7). Clearly, lung transplant recipients urgently need strategies to prevent or delay the onset of CLAD to improve survival after transplantation.

The exact pathogenesis of the different phenotypes of CLAD remains unclear. Studies have consistently identified multiple clinical risk factors including primary graft dysfunction (PGD), acute cellular rejection (ACR), lymphocytic bronchiolitis (LB), and the development of donor-specific human leukocyte antigens (HLA) antibodies (DSA) (2, 8-12). DSA appear to play a central role in the development of CLAD. Studies have consistently identified the development of DSA as an independent risk factor for the development of CLAD (11, 12). Furthermore, DSA cause antibody-mediated rejection (AMR) and are associated with an increased risk of high grade and recurrent ACR as well as LB (13, 14). In addition, DSA may mediate the increased risk of CLAD associated with PGD (15). Lastly, experimental data suggest that DSA have a direct pathogenic role in the development of CLAD (16-18). These data are consistent with the experience with DSA in kidney transplantation (19). However, the development of DSA after lung transplantation is significantly more frequent than after kidney transplantation. Indeed, single center studies have reported an incidence of DSA after lung transplantation ranging between 25-50%, and an incidence of 39% over the first 120 days after transplantation was reported in a recent prospective multicenter study (11, 12, 20). Together, these findings demonstrate that DSA are common early after lung transplantation and are associated with an increased risk of ACR, LB, AMR and CLAD.

We hypothesize that Belatacept-based immunosuppression (IS) will result in a lower incidence of DSA development and improved clinical outcomes after lung transplantation. To test this hypothesis, we plan to conduct a multicenter phase III randomized-controlled trial (RCT) examining the efficacy and safety of Belatacept-based immunosuppression after lung transplantation with a primary endpoint of CLAD-free survival. However, because experience with Belatacept in lung transplantation is very limited, we plan to conduct a pilot 2-center phase II RCT to assess the feasibility of conducting the phase III RCT.

##### 2.3. POTENTIAL RISKS AND BENEFITS

###### 2.3.1. KNOWN POTENTIAL RISKS

Some studies in kidney transplantation have demonstrated an increased risk of ACR among patients treated with Belatacept in the place of a calcineurin inhibitor (CNI) (21, 22). There have been no controlled studies of Belatacept in lung transplantation, but since the risk of ACR after lung transplantation is significantly higher than after kidney transplantation, there may be an increased risk of ACR for those randomized to the Belatacept arm of this study. In addition, Belatacept will be given after transplant surgery on day 0 in this study whereas it had been given pre-operatively or intra-operatively in previous kidney transplant studies, and this may impact its efficacy. We have designed the study protocol to mitigate the potential risk of ACR. First, all study participants will be treated with rabbit anti-thymocyte globulin (ATG, Thymoglobulin®) for induction IS. This is considered a more intensive immunosuppressive induction agent than basiliximab (Simulect®) and clearly more intensive IS than no induction therapy. At the 2 study sites, rabbit ATG is used for induction immunosuppression for patients deemed to be at increased immunologic risk (e.g., those who have DSA at the time of transplant or who have historical DSA) and for treatment of persistent ACR and CLAD. The use of ATG carries potential risks of infusion reaction and hematologic toxicities (leukopenia and thrombocytopenia). To mitigate the risk of infusion reactions, we plan to pre-medicate doses of ATG with methylprednisolone, diphenhydramine, acetaminophen, and an H2 blocker. These pre-medications may be associated with side effects. Typically, these are minor and self-limited, including symptoms such as drowsiness, dry mouth, euphoria, insomnia, increased appetite, and nausea. We will monitor for the development of hematologic toxicities with complete blood counts (CBC) during therapy, and modify subsequent ATG doses accordingly (section **7.4.**). Although the use of ATG for induction immunosuppression after lung transplantation has not been associated with a significantly increased risk of infection in 2 previous randomized controlled trials (RCT) (23, 24), we will use prophylactic antimicrobials (section **7.4.**) and monitor patients for the development of infection with blood testing and bronchoscopy with bronchoalveolar lavage (BAL) (section **7.2.** and **7.3.**). Co-administration of Belatacept and ATG has been associated with an increased risk of venous thrombosis in kidney transplant studies particularly when the 2 drugs are administered in close temporal proximity. To mitigate this risk, we plan to administer the 2 drugs at least 12 hours apart (**7.4.**).

To further mitigate the risk of ACR, participants randomized to the Belatacept arm will be treated with the combination of Belatacept and Tacrolimus for the first 3 months after transplantation. This time period after transplantation carries the highest risk of ACR. We have also implemented 1 additional bronchoscopy with transbronchial lung biopsies at day 252 (± 14) (week 36) to increase surveillance for ACR. Our routine clinical practice at the 2 sites is to perform 5 surveillance bronchoscopies with BAL and transbronchial lung biopsies during the first year after transplantation at approximately the following time points: 30, 60, 90, 180, and 365 days. The study protocol deviates from this clinical practice by changing the time points to: 28, 84, 112, 168, and 365 (± 14) days and adding a procedure on day 252 ± 14 days. Potential risks of an additional bronchoscopy with BAL and transbronchial lung biopsies include pneumothorax, bleeding, and over-sedation. The risk of pneumothorax is approximately 2%. All bronchoscopies are performed under fluoroscopic guidance to minimize this risk. We routinely perform a chest x-ray after the bronchoscopy to evaluate for a pneumothorax. Most cases of post-bronchoscopy pneumothorax are managed conservatively with serial chest x-rays, and less than half of cases require chest tube drainage. The risk of bleeding during bronchoscopy is 2-3%, and this typically resolves spontaneously with natural hemostasis and bronchoscopic aspiration of clots. Rarely, ice-cold saline or topical epinephrine is used to induce mucosal vasoconstriction and expedite hemostasis. Our sites' routine approach to minimize the risk of bronchoscopy-associated bleeding is to ensure appropriate coagulation factors and that the patient is not on an anticoagulant before the procedure. Over-sedation may occur in 1-2% of bronchoscopy procedures. This may manifest with hypotension or hypoxemia; these are typically transient and resolve with conservative measures. Our routine approach to sedation-related hypotension is to administer intravenous fluids. In addition, patients are routinely intubated endotracheally or given high-flow supplemental oxygen during bronchoscopy procedures; this minimizes the risk of hypoxemia related to sedation.

Another potential risk associated with Belatacept-based immunosuppression is post-transplant lymphoproliferative disease (PTLD) (22). This increased risk has been identified specifically among Epstein-Barr Virus (EBV) seronegative recipients of organs from seropositive donors. To minimize the risk of PTLD, we will exclude EBV seronegative patients from this study. In previous RCTs in kidney transplantation, there was no significant difference in the incidence of serious infection or cancer in Belatacept-treated patients compared to those treated with CNIs.

Study participants will undergo 2 additional (blood) DSA tests than the routine clinical protocol at the sites (section **7.3.**) on days 112 and 252 (± 14). These blood samples will be obtained at time points when patients routinely undergo blood draws for clinical purposes (CBC, Tacrolimus trough level) and will pose only minimal additional risk by increasing the volume of blood collected by 5 mL.

##### 2.3.2. KNOWN POTENTIAL BENEFITS

Belatacept-based IS has been consistently associated with better kidney function than CNI-based therapy (21, 22, 25). We expect participants randomized to Belatacept in this study to derive a similar benefit in kidney function. Furthermore, Belatacept-treated kidney transplant recipients appear to be less likely to develop DSA than CNI-treated recipients (25). The development of DSA after lung transplantation is significantly more frequent than after kidney transplantation, and DSA have been linked to all forms of lung allograft rejection. Thus, this potential benefit of Belatacept-based IS may result in better outcomes after lung transplantation.

In kidney transplantation, Belatacept has been associated with a lower incidence of diabetes mellitus, systemic hypertension, and hypercholesterolemia compared to CNIs. Thus, patients randomized to Belatacept in this study may derive similar benefits with a lower risk of these post-transplant metabolic complications. Additionally, patients randomized to Belatacept will not require frequent blood draws for therapeutic Tacrolimus monitoring after day 90 post-transplant.

#### 3. OBJECTIVES AND PURPOSE

The primary objective of this pilot RCT is to assess the feasibility of conducting a phase III multicenter RCT examining the efficacy and safety of Belatacept-based immunosuppression after lung transplantation.

This pilot study may identify potential problem areas in the current protocol that require modification to improve feasibility. For example, eligibility criteria may be modified in a future RCT based on experience in this pilot study. Similarly, concomitant or prophylactic medications may be modified to improve protocol adherence or participant retention in the future RCT.

#### 4. STUDY DESIGN AND ENDPOINTS

##### 4.1. STUDY DESIGN

This is a pilot 2-center open-label RCT comparing Belatacept-based immunosuppression to standard of care immunosuppression consisting of the combination of Tacrolimus, Mycophenolate Mofetil (MMF), and Prednisone.

##### 4.2.1. PRIMARY ENDPOINT

The primary endpoint of the study is the feasibility metric of randomizing 80% of patients eligible for randomization within 4 hours of completion of transplantation.

##### 4.2.2. SECONDARY ENDPOINTS

The following secondary endpoints are chosen because they include important objective metrics of feasibility and they have a clinical impact on outcomes after lung transplantation. Clinical secondary endpoints (**c.** – **z.** below) will be assessed 1 year after transplantation. These will be used to inform the design of the future phase III RCT and will be important in making appropriate power and sample size calculations.

- **a.** Enrollment of 50% of patients eligible for enrollment at the 2 centers
- **b.** Retention of 75% of randomized patients on the protocol
- **c.** Development of DSA with mean fluorescence intensity (MFI) ≥ 2000
- **d.** Death
- **e.** Re-transplantation
- **f.** ACR International Society for Heart and Lung Transplantation (ISHLT) grade A1 or higher
- **g.** ACR ISHLT grade A2 or higher
- **h.** LB ISHLT grade B1R or higher
- **i.** DSA with MFI ≥ 4000
- **j.** DSA Immunoglobulin G (IgG) subclasses
- **k.** Complement binding (C1q-positive) and activating (C3d) DSA
- **l.** Definite AMR based on the ISHLT definition
- **m.** Probable AMR based on the ISHLT definition
- **n.** CLAD defined as BOS stage 1 or RAS
- **o.** CLAD-free survival
- **p.** Confirmed bacterial infection requiring antibiotic treatment
- **q.** Cytomegalovirus (CMV) infection requiring antiviral treatment
- **r.** Confirmed community-acquired respiratory viral infection (CARV)
- **s.** Chronic kidney disease (CKD) stage 3 as estimated by the Cockcroft-Gault equation
- **t.** Kidney function as estimated by the Cockcroft-Gault equation at 6 months and 12 months after randomization
- **u.** Malignancy excluding squamous cell and basal cell skin cancer
- **v.** Post-transplant Lymphoproliferative Disease (PTLD)
- **w.** Progressive multifocal leukoencephalopathy
- **x.** Diabetes mellitus requiring medical treatment
- **y.** Systemic hypertension requiring medical treatment
- **z.** Hypercholesterolemia requiring medical treatment

##### 4.3. PROGRESSION CRITERIA TO PHASE III RCT

The decision to proceed to a phase III multicenter RCT will be based on the totality of results from this pilot study. It is possible that we may proceed with the phase III RCT even if the primary endpoint in this pilot study is not met. For example, we may identify certain barriers to randomization in this pilot study that are modifiable in the phase III RCT. Additionally, we will consider the other objective assessments of feasibility (Secondary Endpoints **a.** and **b.**) and the clinical efficacy endpoints (Secondary Endpoints) in determining progression to the phase III RCT.

#### 5. STUDY ENROLLMENT AND WITHDRAWAL

We will enroll patients after listing for transplantation according to the eligibility criteria below (**5.1.** and **5.2.**). During transplant surgery, some patients may develop complications that place them at increased risk of poor outcomes (e.g., severe PGD requiring ECMO support). We believe that such patients are best managed according to the sites' routine clinical protocols rather than be randomized in this study and potentially be treated with an investigational immunosuppressive regimen. Therefore, we will assess enrolled patients' eligibility for randomization and randomize them if eligible within 4 hours of completion of transplantation (**7.1.**). Below is a schematic of the timeline of enrollment and randomization:

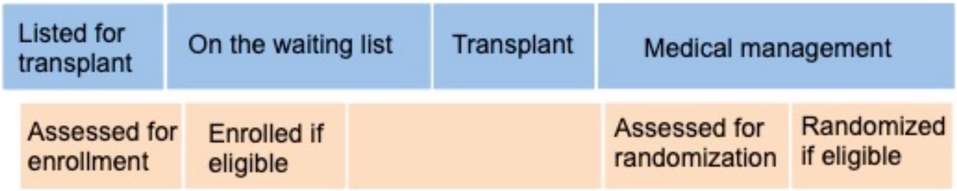


##### 5.1. INCLUSION CRITERIA FOR ENROLLMENT

- **a.** Adult between 18 to 70 years of age
- **b.** On the waiting list for lung transplantation
- **c.** Capable of understanding the purposes and risks of the study in English and willing to participate and sign informed consent
- **d.** For women of child bearing potential, willing to use highly effective contraception
- **e.** Willing to return to the medical center every 4 weeks for care after week 12 post-transplant

##### 5.2. EXCLUSION CRITERIA FOR ENROLLMENT

Individuals who meet any of the following exclusion criteria will be excluded from study participation:

- **a.** Requiring invasive mechanical ventilation immediately before transplantation
- **b.** Requiring extracorporeal life support (ECLS) (i.e., ECMO) immediately before transplantation
- **c.** Received treatment to deplete HLA antibodies before transplantation
- **d.** Having DSA immediately before transplantation (i.e., positive virtual crossmatch)
- **e.** Multi-organ transplant recipient (e.g., heart-lung, lung-liver, lung-kidney)
- **f.** Pregnant or breast-feeding
- **g.** Active infection with Hepatitis B virus
- **h.** Active infection with Hepatitis C virus
- **i.** Active infection with human immunodeficiency virus (HIV)
- **j.** Chronic infection with *Burkholderia cepacia* complex before transplantation
- **k.** EBV seronegative status
- **l.** Participation in another interventional clinical trial
- **m.** Any condition that in the opinion of the site PI introduces undue risk by participating in this study
- **n.** Incapable of understanding the purposes of the study or informed consent due to a language barrier or cognitive deficits

##### 5.3. STRATEGIES FOR RECRUITMENT AND RETENTION

We will recruit study participants from the 2 sites' lung transplant clinics or inpatient services (if appropriate candidates are hospitalized at the time of listing). The investigators or the study research coordinators will screen and approach potential participants after listing for transplantation. We will explain the study and the potential benefits and risks involved in participation to patients during the informed consent process. We will specifically explain that participation is entirely voluntary and will not impact patients' waiting time or their likelihood to receive a transplant. We will verbally re-affirm enrolled patients' assent to participate in this study by telephone monthly while on the waiting list and note this in their study files. The study will not enroll children (< 18 years of age) and will not enroll vulnerable participants (i.e., those who lack the capacity to consent or those who may perceive coercion to participate) or those who do not understand English. We recognize the importance of minorities and women in clinical research, and we will not exclude participants from this study based on race or gender. Race and gender characteristics of patients being listed for transplantation are proportionate to the underlying diagnoses and referral patterns. A targeted/planned enrollment table is shown below:

| | Ethnic categories | | | | | | |
|---|---|---|---|---|---|---|---|
| Racial categories | Not Hispanic or Latino | | | Hispanic or Latino | | | |
| | Female | Male | Unknown | Female | Male | Unknown | Total |
| American Indian | 0 | 0 | 0 | 0 | 0 | 0 | 0 |
| Asian | 2 | 2 | 0 | 0 | 0 | 0 | 4 |
| Pacific Islander | 0 | 0 | 0 | 0 | 0 | 0 | 0 |
| Black | 5 | 6 | 0 | 0 | 0 | 0 | 11 |
| White | 24 | 28 | 0 | 7 | 8 | 0 | 67 |
| More than 1 race | 0 | 0 | 0 | 0 | 2 | 0 | 2 |
| Unknown | 0 | 0 | 0 | 0 | 0 | 0 | 0 |
| Total | 31 | 36 | 0 | 7 | 10 | 0 | 84 |

Some enrolled patients may not undergo transplantation during the study period. Other enrolled patients who undergo transplantation may not be eligible for randomization. We estimate that 60% of enrolled patients will be eligible for randomization. We plan to enroll 84 patients and estimate that 50 (84 * 0.6) will be eligible for randomization, and 40 (80%) will be randomized (50 * 0.8). Over the past 3 years, a mean 150 patients have undergone lung transplantation annually at the 2 sites. Based on these data, we expect that approximately 187 patients will undergo transplantation at the 2 sites over the 15-months study enrollment period. We have planned to enroll a conservative number of patients but plan to screen all patients listed for transplantation at the 2 sites and approach those who are eligible to consent. We plan to compensate study participants with gift cards valued $130 to improve retention.

##### 5.4. PARTICIPANT WITHDRAWAL OR STUDY DRUG DISCONTINUATION

Participants are free to withdraw from study participation at any time upon request. The investigators may discontinue the study drug (Belatacept) temporarily or permanently if a participant has an adverse event (AE), a serious adverse event (SAE), or any medical condition such that continuation of the study drug would not be in the best interest of the participant. Upon discontinuation of the study drug, these participants will be treated with a regimen deemed most appropriate by the patient's primary transplant physician but will continue to be followed by the study for safety and efficacy endpoints. The investigators may discontinue the study drug for any of the indications outlined in **5.4.1.**:

##### 5.4.1. INDIVIDUAL PATIENT-SPECIFIC STOPPING RULES

a. Three episodes of ACR grade A2 or higher
b. Three episodes of LB grade B1R or higher
c. Development of CLAD
d. Development of AMR
e. Serious infectious complication that in the opinion of investigator warrants changing the maintenance immunosuppressive regimen
f. Malignancy including PTLD, but excluding skin cancer

##### 5.5. STUDY-WIDE STOPPING RULES

Circumstances that may warrant termination or suspension of the study include determination of unexpected, significant, or unacceptable risk to participants. Study-wide stopping rules will be based on the comparison of rates of the following clinical outcomes between the 2 treatment groups:

a. Incidence of DSA with MFI ≥ 4000 or C1q-positive DSA
b. Incidence of CLAD
c. Incidence of AMR
d. Incidence of serious infection that in the opinion of the investigator warrants changing the maintenance immunosuppressive regimen
e. Incidence of PGD grade 3 at 72 hours after transplantation
f. Need for ECMO support within 7 days of transplantation
g. Listing for re-transplantation
h. Death

The Data Coordinating Center (DCC) will tabulate and compare the rate of development of the above outcomes (**5.5.**a.-h.) between the 2 treatment groups after every 10 patients are randomized and have had at least 6 months of follow-up and report the results to the investigators and the DSMB. After the first 10 participants have completed 6 months of follow-up, subsequent reports will include all data from all participants who have completed at least 6 months of follow-up and all randomized participants with available data at that time. Based on these results, the DSMB may suspend or terminate the study. If the incidence of any of the above outcomes (**5.5.**a.-h.) in the Belatacept arm is greater than 50%, and this incidence is more than 50% higher than the incidence in the standard of care immunosuppression arm, the investigators will seek guidance from the DSMB about study continuation, suspension, or termination.

The DSMB may develop additional stopping rules for safety concerns. Safety data including AEs, SAEs, laboratory test abnormalities, and treatment failures will be presented by treatment group at each DSMB meeting. Summary statistics and/or plots of the key outcomes will be provided to the DSMB at each meeting to allow appropriate assessment of the relative risks and benefits. We do not plan to include a futility monitoring plan or an efficacy-stopping rule because this is a phase II trial with the potential for collecting valuable information for designing the subsequent clinical trial or identifying potential subgroups of interest. The investigators will promptly inform the IRB, Bristol-Myers Squibb (BMS), and the NIH and provide the reasons for temporary suspension or termination. If suspended, the study may resume once concerns about safety, protocol compliance, or data quality are addressed and satisfy the necessary regulatory agencies.

**5.6. ETHICAL CONSIDERATIONS**

The study will be conducted in the following manner:

a. In accordance with Good Clinical Practice (GCP), as defined by the International Conference on Harmonization (ICH), World Health Organization (WHO) and any local directives.
b. In compliance with the protocol.
c. The protocol, any amendments, and the subject informed consent will receive Institutional Review Board / Independent Ethics Committee (IRB/IEC) approval / favorable opinion before initiation of the study.
d. With personnel who are qualified by education, training, and experience to perform their respective tasks and that the study will not use the services of study personnel for whom sanctions have been invoked or where there has been scientific misconduct or fraud.
e. With signed, dated informed consent from each of the participants.
f. Investigators must ensure that subjects are clearly and fully informed about the purpose, potential risks, and other critical issues regarding clinical studies in which they volunteer to participate. The approved informed consent form will adhere to the ethical principles that have their origin in the Declaration of Helsinki.
g. Inclusion of relevant safety information regarding dose / schedule of IP and any other drugs / procedures.
h. BMS and health authorities will have direct access to study records.
i. Inclusion of information outlining BMS support of the study (study drug, funding, etc.)

**6. STUDY DRUG**

Belatacept (Nulojix) will be provided by BMS from commercial supply for use in this study. Lot numbers will be assigned at study initiation. BMS will review and approve the study protocol and receive regular progress reports documenting enrollment and randomization. Additionally, BMS will receive safety data in real time (section **8.3.**). Belatacept is a lyophilized powder and is supplied as a single use 250 mg vial with one 12 ml silicone free disposable syringe. Belatacept will be shipped directly from BMS to Washington University School of Medicine in St. Louis and Houston Methodist Hospital and will be stored and dispensed by the sites' investigational drug service pharmacies. Belatacept must be stored refrigerated at 2°to 8°C (36°-46°F) and protected from light by storing in the original package until time of use per package insert. For details, see the Belatacept (Nulojix) package insert and Investigator's Brochure. Belatacept will be clearly labeled as being for research purposes only.

An Investigational Drug Accountability Log entry must be completed by the sites' investigational pharmacist and emailed to the Clinical Coordinating Center (Washington University Sponsor-Investigator) each time Belatacept is received, dispensed, or destroyed. All expired or unused Belatacept and empty containers at the sites can be disposed of per the institution's standard drug destruction procedure after notifying BMS. Written document of what vials were discarded, and the date is mandatory. (Otherwise all expired drug must be quarantine and sent back to the company). A daily temperature log describing maximum and minimum daily temperature of refrigerated storage will also be required.

Belatacept will be dosed at 10 mg/kg on days 0, 7, 14, 28, 56, 84, then at 5 mg/kg on days 112, 140, 168, 196, 224, 252, 280, 308, 336 and 364, where day 0 is the day of transplant. The first dose of Belatacept will be given after transplant surgery. This is different from previous studies in kidney transplantation where the first dose was given pre-operatively or intra-operatively. Although this may impact its efficacy, administering Belatacept after transplant surgery is necessary in this protocol to avoid including recipients who develop serious complications intra-operatively that place them at increased risk of poor outcomes in this small pilot study (**7.1.2.**). After the initial 4 doses on days 0, 7, 14 and 28, subsequent doses may be given ± 3 days. Belatacept will be given intravenously over 30 minutes. During the index hospitalization, Belatacept will be given in the patient's hospital room; after discharge from the hospital, Belatacept will be given at the sites' outpatient infusion center.

#### 7. STUDY PROCEDURES AND SCHEDULE

##### 7.1. RANDOMIZATION

We will assess enrolled patients' eligibility for randomization within 4 hours of completion of transplantation (transfer to the intensive care unit from the operating room).

###### 7.1.1. INCLUSION CRITERIA FOR RANDOMIZATION

- **a.** Provided written informed consent for participation in this study
- **b.** Underwent single or bilateral lung transplant
- **c.** Negative urine pregnancy test for women of child bearing potential

###### 7.1.2. EXCLUSION CRITERIA FOR RANDOMIZATION

- **a.** Allograft dysfunction requiring ECMO support
- **b.** Delayed chest closure (i.e., primary chest closure not yet performed)
- **c.** Severe coagulopathy and clinically significant bleeding in the opinion of the site PI
- **d.** Any condition that in the opinion of the site PI introduces undue risk by participating in this study

###### 7.1.3. RANDOMIZATION PROCEDURE AND GROUPS

Enrolled participants will be assessed within 4 hours of arriving in the ICU after transplant. Eligible participants will be randomized, within center, and blocked to prevent temporal bias using a computer-generated method (REDCap [Research Electronic Data Capture] randomization module) with a 1:1 ratio to on one of 2 treatment assignments:

- **a.** Belatacept-based immunosuppression
  - **i.** Belatacept + Tacrolimus + Prednisone from day 0 through day 89, then
  - **ii.** Belatacept + Mycophenolate Mofetil + Prednisone from day 90 through day 365
- **b.** Tacrolimus + Mycophenolate Mofetil + Prednisone from day 0 through day 365

At the end of year 1 post-transplant, patients randomized to Belatacept will resume Tacrolimus on day 392 (28 days of the last Belatacept dose) and continue Mycophenolate Mofetil and Prednisone.

##### 7.1.4. BELATACEPT DOSING AND DIAGNOSTIC TESTING SCHEDULE

| | BELA 10 mg/kg<br>Tacrolimus<br>Prednisone | BELA 5 mg/kg<br>MMF / MPA<br>Prednisone |
|---|---|---|
| BELA | D0, D7, D14, W4, W8, W12 | W16, W20, W24, W28, W32, W36, W40, W44, W48, W52 |
| TBBX | W4, W12 | W16, W24, W36, W52 |
| DSA | D0, D10, W4, W8, W12 | W16, W24, W36, W52 |
| CIP | D0, D10, W4, W8, W12 | W16, W24, W36, W52 |

#### 7.2. STUDY AND STANDARD OF CARE PROCEDURES

Routine clinical assessment of study participants will be performed according to the sites' clinical protocols. After discharge from the transplant hospitalization, patients are seen in the lung transplant clinic every 1-2 weeks for the first 3 months after transplantation then every 4 weeks through the 1st year after transplantation at both sites. These clinic visits include a medical history, medication history, physical examination, routine lab work, chest x-ray, and spirometry. Certified and trained respiratory therapists will conduct spirometry measurements according to American Thoracic Society (ATS) guidelines. Unscheduled visits are arranged on an as-needed basis if patients develop signs or symptoms of allograft dysfunction or another complicating condition.

Participants will undergo fiberoptic bronchoscopy (FOB) with bronchoalveolar lavage (BAL) and transbronchial lung biopsies (TBBX) on days 28, 84, 112, 168, 252, and 365 (± 14 days). As noted above (section **2.3.1.**), this bronchoscopy schedule is modified from the clinical protocol and the day 252 procedure was included to provide an additional screening time point for ACR. In addition, patients will undergo bronchoscopy with BAL and lung biopsies for clinical indications (i.e., if they develop signs or symptoms of allograft dysfunction) and 3-6 weeks after an episode of ACR to exclude persistent ACR according to the sites' clinical protocols. The sites' clinical pathologists will interpret the lung biopsy results and report these to the patients' clinicians as is routinely done in clinical practice. However, the Washington University pathology lab will serve as the Study Pathology Core Lab, and the Study Pathology Core Lab Investigator (Dr. Gaut) will interpret all biopsies. Biopsies from Houston Methodist Hospital will be shipped to the Study Pathology Core Lab. Dr. Gaut will be blinded to the participants' treatment assignment and study visit. Only the core lab's pathology interpretation will be analyzed for study purposes.

#### 7.3. LABORATORY PROCEDURES AND EVALUATIONS

Routine blood work including complete blood counts (CBC), comprehensive metabolic panels (CMP), blood CMV polymerase chain reaction (PCR), and trough levels of tacrolimus will be done according to the sites' clinical protocols 1-2 times weekly during the first 3 months after transplantation then monthly through the 1st year after transplantation. In between these time points, patients may require unscheduled lab evaluations on an as needed basis. A urine pregnancy test will be performed on all women of child bearing potential at enrollment, at randomization, and before conversion from Tacrolimus to mycophenolate mofetil on day 90 for study participants randomized to Belatacept therapy.

All lung transplant candidates undergo HLA typing and SAB testing for HLA antibodies prior to listing. Donors are HLA typed and the information is available when organs are offered for transplant. These data are used to perform a virtual crossmatch prior to transplant. Flow cytometry crossmatch using donor tissue and recipient blood is performed after transplant. DSA will be defined as a donor-specific HLA antibody with a mean fluorescence intensity (MFI) ≥ 2000. Participants' serum will be tested for DSA on days 0 and 10 (± 3 days), and days 28, 56, 84, 112, 168, 252, and 365 (± 14 days). This DSA testing schedule coincides with the study bronchoscopy and lung biopsy schedule and is a modified version of the sites' clinical DSA testing protocol (days 0, 10, 30, 60, 90, 180, 365 ± 14 days). Samples will be tested for DSA using Luminex Single Antigen Bead (SAB) assay (One Lambda, Canoga Park, CA). If a sample tests positive for DSA, follow up testing will be performed for IgG subclasses (IgG 1, 2, 3 and 4), and markers of complement binding and activation (C1q and C3d). The Baylor University Medical Center (BUMC) Transplant Immunology Lab will serve as the study core lab for antibody testing. DSA testing will also be conducted at the sites' HLA labs and the results will be reported to the patients' clinicians as is routinely done in clinical practice. However, only results from the study core lab will be analyzed for study purposes. Specimens from study participants will shipped to the BUMC core lab for processing and analysis. To reduce potential bias related to the study's open label design, the HLA investigator (Dr. Askar) and all laboratory personnel will be blinded to the participants' treatment assignment and study visit. Blood samples will be drawn at the same time points and stored for future cellular immunophenotyping (CIP) assays using pre-validated DuraClone IM flow cytometry panels (Beckman Coulter, Brea, CA) (26). These assays will enable longitudinal characterization of peripheral blood mononuclear cell (PBMC), B, T and T regulatory cell populations under the two treatment conditions. In addition, samples will be stored in Streck BCT tubes for future cell free DNA analysis.

Study participants of child bearing potential will have urine pregnancy tests at enrollment, randomization, and at the time of initiation of Mycohphenolate Mofetil for those randomized to Belatacept.

##### 7.4. CONCOMITANT AND PROPHYLACTIC MEDICATIONS

All study participants will be treated with ATG (1 mg/kg on post-operative day 0, 1 and 2, cumulative dose 3 mg/kg) for induction immunosuppression. ATG will be given intravenously over 6-8 hours in the patient's hospital room. The first dose of ATG will be given at randomization. For participants randomized to the Belatacept arm, the first dose of Belatacept will given 12 hours after the first dose of ATG is given. Subsequent ATG dosing may be modified in the event of hematologic toxicity (i.e., white blood cell (WBC) count < 4.0 K/mm$^3$ or platelet count < 70 K/mm$^3$). Subsequently, the second dose of ATG will be given 12 hours after Belatacept. Tacrolimus will be initiated enterally or sublingually within the first 48 hours after transplantation and dosed to target a trough blood level of 8-15 ng/mL if kidney function is normal. In case of abnormal kidney function, target Tacrolimus trough blood levels will be decreased accordingly to 4-8 ng/mL. On day 90, mycophenolate mofetil (MMF) will be substituted for tacrolimus in the Belatacept arm and will be dosed at 1 g twice daily (or converted to enteric coated mycophenolic acid (Myfortic) 720 mg twice daily in the event of gastrointestinal toxicity). In the control arm, MMF will be initiated intravenously at 1 g twice daily on day 0 and will be converted to enteral dosing at 1 g twice daily once the patient can take oral medications or enteric coated mycophenolic acid (Myfortic) 720 mg twice daily in the event of gastrointestinal toxicity. In the event of persistent gastrointestinal toxicity in spite of changing to enteric coated mycophenolic acid, patients will be treated with azathioprine 2 mg/kg daily. All study participants will be treated with methylprednisolone 500 mg intravenously before perfusion of the allograft during the transplant procedure, then methylprednisolone 0.5 mg/kg intravenously twice daily for 6 doses, then prednisone 0.5 mg/kg orally daily through day 14, then 0.2 mg/kg daily through day 30, then 0.1 mg/kg daily through day 180, then 5 mg daily through day 365.

All study participants will be treated with trimethoprim/sulfamethoxazole (Bactrim), dapsone, inhaled pentamidine, or atovaquone for *Pneumocystis jirovecii* prophylaxis throughout the study period. Recipients at risk for CMV infection (i.e., seronegative recipients of seropositive donors or seropositive recipients) will be treated with valganciclovir for CMV prophylaxis through day 365. Recipients who are not at risk for CMV infection (i.e., seronegative recipients of seronegative donors) will be treated with acyclovir or valacyclovir for herpes simplex virus (HSV) and varicella zoster virus (VZV) prophylaxis throughout the study period. Antifungal prophylaxis will be tailored to culture results from bronchoscopy specimens and pre-transplant cultures.

#### 7.5. CLINICAL MANAGEMENT OF POST-TRANSPLANT COMPLICATIONS

This section provides an overview of our sites' clinical protocols for the management of common complications after lung transplantation and is not intended to be comprehensive. It is possible that there may be discordance between the sites' local pathology or DSA interpretation and the study's core lab results. The decision to initiate treatment for ACR or DSA will be based on the sites' local pathology or HLA lab's results, but the study's core lab results will be reported to the sites' PIs as soon as they are available. Episodes of ACR grade A2 or higher are treated with methylprednisolone 500 mg intravenously daily for 3 days. The decision to initiate treatment for episodes of ACR grade A1 is individualized based on the patient's previous history of ACR, infection, and lung function. A follow-up bronchoscopy with BAL and lung biopsies is performed 3-6 weeks after an episode of ACR to exclude persistent ACR. The decision to initiate treatment for episodes of LB is individualized based on the patient's previous history of ACR, LB, infection, concomitant bronchoscopy culture results, and lung function. Similarly, according to the sites' clinical protocols, the decision to initiate treatment and the choice of treatment for DSA is individualized based on the patient's history of ACR, DSA characteristics (e.g., HLA Class, DSA locus, MFI), lung function, and history of infection.

Drug-induced leukopenia (WBC count < 4.0 K/mm$^3$) due to MMF (or mycophenolic acid) alone or in combination with valganciclovir is managed by first withholding MMF (or mycophenolic acid) until the WBC count normalizes. Persistent and more severe leukopenia (WBC count < 2.5 K/mm$^3$) in spite of withholding MMF is managed by then withholding valganciclovir.

Cases of CMV infection (isolated viremia or viremia with invasive disease) are managed with either oral valganciclovir or intravenous ganciclovir as clinically appropriate. Confirmed or suspected bacterial infections are managed with oral or intravenous antibiotics as clinically appropriate. The management of CARV infections is individualized depending on the specific virus and the presence and severity of lower respiratory tract symptoms. For example, respiratory syncytial virus (RSV) infection is generally treated with ribavirin whereas parainfluenza virus (PIV) and rhinovirus infection are treated with supportive care. Lower respiratory tract symptoms such as wheezing, cough, or a significant decline in lung function are managed with high-dose steroids and empiric antibacterial antibiotics for a possible superimposed bacterial infection. All patients with infectious symptoms are followed closely until resolution or improvement of these symptoms. In all cases of severe infection, if in the opinion of the investigator or the patient's transplant clinician changing the maintenance immunosuppressive regimen is warranted, the patient will be withdrawn from the study protocol and managed as deemed clinically appropriate (section **5.4.1.**).

#### 7.6. SCHEDULE OF EVENTS TABLE

Below is a table illustrating the schedule of study and clinical events. Time windows for each time point are listed above. Unscheduled visits may occur at any time point after randomization, and data from these unscheduled visits will be included in the study analyses as appropriate.

| TIME POINTS | | | | | | | | | | | | | | | | | |
|---|---|---|---|---|---|---|---|---|---|---|---|---|---|---|---|---|---|
| Week | | 0 | | 4 | 8 | 12 | 16 | 20 | 24 | 28 | 32 | 36 | 40 | 44 | 48 | 52 | 56 |
| Day | | 0 | 10 | 28 | 56 | 84 | 112 | 140 | 168 | 196 | 224 | 252 | 280 | 308 | 336 | 365 | 395 |
| **EVENTS** | | | | | | | | | | | | | | | | | |
| Screening | X | | | | | | | | | | | | | | | | |
| Informed consent | X | | | | | | | | | | | | | | | | |
| Medical history | X | X | | | | | | | | | | | | | | | |
| Pregnancy test | X | X | | | | X | | | | | | | | | | | |
| Physical exam | X | X | X | X | X | X | X | X | X | X | X | X | X | X | X | X | X |
| Randomization | | X | | | | | | | | | | | | | | | |
| CBC, CMP | | | X | X | X | X | X | X | X | X | X | X | X | X | X | X | X |
| Drug level | | | X | X | X | X | X | X | X | X | X | X | X | X | X | X | X |
| CMV blood PCR | | | X | X | X | X | X | X | X | X | X | X | X | X | X | X | X |
| Chest X-ray | | X | X | X | X | X | X | X | X | X | X | X | X | X | X | X | X |
| Spirometry | | | | X | X | X | X | X | X | X | X | X | X | X | X | X | |
| DSA | | X | X | X | X | X | X | | X | | | X | | | | X | |
| CIP | | X | X | X | X | X | X | | X | | | X | | | | X | |
| TBBX | | | | X | | X | X | | X | | | X | | | | X | |
| AE evaluation | | | X | X | X | X | X | X | X | X | X | X | X | X | X | X | X |

#### 8. SAFETY CONSIDERATIONS

The study safety reporting period commences once a patient provides written informed consent to participate in the study and will be complete through day 395. This includes 30 days of follow-up after the last dose of Belatacept in the group randomized to Belatacept.

#### 8.1. SERIOUS ADVERSE EVENT DEFINITION

A ***Serious Adverse Event (SAE)*** is any untoward medical occurrence that at any dose:

a. Results in death
b. Is life-threatening (defined as an event in which the subject was at risk of death at the time of the event; it does not refer to an event which hypothetically might have caused death if it were more severe)
c. Requires inpatient hospitalization or causes prolongation of existing hospitalization
d. Results in persistent or significant disability or incapacity
e. Results in a congenital anomaly or birth defect
f. Is an important medical event (defined as a medical event(s) that may not be immediately life-threatening or result in death or hospitalization but, based upon appropriate medical and scientific judgment, may jeopardize the subject or may require medical or surgical intervention to prevent one of the other serious outcomes listed in the definitions above. Examples of such events include, but are not limited to, intensive treatment in an emergency department or at home for allergic bronchospasm; blood dyscrasias or convulsions that do not result in hospitalization.
g. Results in potential Drug Induced Liver Injury (DILI)
h. Results in suspected transmission of an infectious agent (pathogenic or nonpathogenic) via the study drug
i. Although pregnancy, overdose and cancer are not always considered serious by regulatory definition, these events must be handled as SAEs.

The following hospitalizations are **not** considered SAEs:

a. A visit to the emergency room or other hospital department < 24 hours, that does not result in admission (unless considered an important medical or life-threatening event)
b. Elective surgery, planned prior to signing consent

c. Admissions as per protocol for planned medical or surgical procedure
d. Routine health assessment requiring admission for baseline/trending of health status (e.g. routine colonoscopy)
e. Medical / surgical admission other than to remedy ill health and planned prior to entry into the study. Appropriate documentation is required in these cases.
f. Admission encountered for another life circumstance that carries no bearing on health status and requires no medical or surgical intervention (e.g. lack of housing, economic inadequacy, caregiver respite, family circumstances, administrative reason.

##### 8.2. SERIOUS ADVERSE EVENT REPORTING – WASHINGTON UNIVERSITY SPONSOR-INVESTIGATOR

Procedures for safety reporting to Washington University Sponsor-Investigator (Clinical Coordinating Center) are described below:

a. All SAEs that occur following a study subject's written consent in the study through 30 days of discontinuation of study drug dosing will be reported to Washington University Sponsor-Investigator through the Data Coordinating Center (DCC) on SAE-specific Case Report Forms through the study's electronic database within 7 days.
b. Following the study subject's written consent to participate in the study, all SAEs, whether related or not related to the study drug, are collected, including those thought to be associated with protocol-specified procedures. The Investigators will report any SAE occurring after these time periods that is believed to be related to study drug or protocol-specified procedure.
c. In accordance with local regulations, Washington University Sponsor-Investigator will notify Investigators of all reported SAEs that are suspected (related to the investigational product) and unexpected (i.e., not previously described in the investigator brochure). An event meeting these criteria is termed a Suspected, Unexpected Serious Adverse Reaction (SUSAR). Investigator notification of these events will be in the form of a SUSAR report.
d. Other important findings which may be reported by Washington University Sponsor-Investigator as an Expedited Safety Report (ESR) include: increased frequency of a clinically significant expected SAE, an SAE considered associated with study procedures that could modify the conduct of the study, lack of efficacy that poses significant hazard to study subjects, clinically significant safety finding from a nonclinical (e.g. animal) study, important safety recommendations from a study data monitoring committee, or Sponsor decision to end or temporarily halt a clinical study for safety reasons.
e. Upon receiving an ESR, the Investigators must review and retain the ESR with the investigator brochure (IB). Where required by local regulations, investigators will submit the ESR to the appropriate IRB. The Investigators and IRB will determine if the informed consent requires revision. The Investigators should also comply with the IRB procedures for reporting any other safety information.
f. Washington University Sponsor-Investigator will report suspected SAEs (whether expected or unexpected) to the relevant local health authorities (either as expedited and/or in aggregate reports).
g. SAEs reported to Washington University Sponsor-Investigator will be tabulated by the DCC, and summary statistics of key outcomes will be provided to the DSMB at each meeting to allow appropriate assessment of the relative risks and benefits. The DCC will also report SAEs to the study steering committee and the FDA.
h. If only limited information is initially available, follow-up reports will be submitted. (Follow-up SAE reports will include the same Investigator term(s) initially reported.)

i. If an ongoing SAE changes in its intensity or relationship to the study drug or if new information becomes available, a follow-up SAE report will be sent within 24 hours to BMS (or designee) using the same procedures used for transmitting the initial SAE report.
j. All SAEs will be followed to resolution or stabilization.

**8.3. SERIOUS ADVERSE EVENT REPORTING – BMS**

Procedures for safety reporting to BMS are described below:

a. All SAEs that occur following a study subject's written consent in the study through 30 days of discontinuation of study drug dosing will be reported to BMS Worldwide Safety: by Washington University Sponsor-Investigator.
b. A MedWatch Form 3500A (Food & Drug Administration) reviewed and approved by BMS will be used to report all SAEs. The BMS protocol ID number will be included on all Form 3500A submitted by the Sponsor or Investigators.
c. Following the study subject's written consent to participate in the study, all SAEs, whether related or not related to the study drug, are collected, including those thought to be associated with protocol-specified procedures. The Investigators will report any SAE occurring after these time periods that is believed to be related to study drug or protocol-specified procedure.
d. SAEs, whether related or not related to study drug, and pregnancies will be reported to BMS within 24 hours. SAEs will be recorded on a BMS approved MedWatch Form 3500A; pregnancies will be reported on a Pregnancy Surveillance Form.

   **SAE FAX Number:** 609-818-3804

e. If only limited information is initially available, follow-up reports will be submitted. (Follow-up SAE reports will include the same Investigator term(s) initially reported.)
f. If an ongoing SAE changes in its intensity or relationship to the study drug or if new information becomes available, a follow-up SAE report will be sent within 24 hours to BMS (or designee) using the same procedures used for transmitting the initial SAE report.
g. All SAEs will be followed to resolution or stabilization.

**8.4. SERIOUS ADVERSE EVENT REPORTING – FOOD & DRUG ADMINISTRATION (FDA)**

Procedures for safety reporting to the FDA are described below:

a. Washington University Sponsor-Investigator will report any event that is both serious and unexpected to the FDA as soon as possible and no later than 7 days (for a death or life-threatening event) or 15 days (for all other Suspected Unexpected Serious Adverse Reactions) after the Investigator or Institution's receipt of the information. BMS will be provided with a simultaneous copy of all adverse events filed with the FDA.
b. SAEs will be reported on MedWatch Form 3500A via electronic submission (http://www.accessdata.fda.gov/scripts/medwatch/). Alternatively, Form 3500A can be sent to the FDA at:

   MEDWATCH
   6500 Fishers Lane
   Rockville, MD 20852-9787
   FAX: 1-800-FDA-0178 (1-800-332-0178)

##### 8.5. SERIOUS ADVERSE EVENT REPORTING – DATA AND SAFETY MONITORING BOARD (DSMB)

The investigators will report the following events to the chair of the DSMB by email:

a. Any event that results in death within 1 working day of the investigators' receipt of the information.
b. Any life-threatening event or unanticipated problem as soon as possible and no later than 7 days after the investigators' receipt of the information.
c. All other Suspected Unexpected Serious Adverse Reactions as soon as possible and no later than 15 days after the investigators' receipt of the information.

##### 8.6. ADVERSE EVENT DEFINITION

An ***Adverse Event (AE)*** is defined as any new untoward medical occurrence or worsening of a pre-existing medical condition in a clinical investigation subject administered an investigational (medicinal) product and that does not necessarily have a causal relationship with this treatment. An AE can therefore be any unfavorable and unintended sign (such as an abnormal laboratory finding), symptom, or disease temporally associated with the use of investigational product, whether or not considered related to the investigational product.

Causal relationship to the study drug is determined by a physician and will be used to assess all adverse events. The causal relationship can be one of the following:

a. Related: There is a reasonable causal relationship between study drug administration and the AE.
b. Not Related: There is not a reasonable causal relationship between study drug administration and the AE.

The term "reasonable causal relationship" means there is evidence to suggest a causal relationship.

Adverse effects can be spontaneously reported or elicited during open-ended questioning, examination, or evaluation of a subject. In order to prevent reporting bias, subjects should not be questioned regarding the specific occurrence of one or more AEs.

##### 8.7. NONSERIOUS ADVERSE EVENT REPORTING

A ***nonserious adverse event*** is defined as an AE not classified as serious

a. Nonserious adverse events will be provided to Washington University Sponsor-Investigator and BMS in aggregate via interim or final study reports as specified in the agreement or as part of an annual reporting requirement (FDA IND).
b. The collection of nonserious AE information will begin at the initiation of the study drug. All nonserious AE (not only those deemed to be treatment-related) will be collected continuously during the treatment period and for a minimum of 30 days after the last dose of study drug.
c. Nonserious AEs will be followed to resolution or stabilization, or reported as SAEs if they become serious. Follow-up will also be conducted for nonserious AEs that cause interruption or discontinuation of study drug and for those present at the end of study treatment as appropriate.

#### 8.8. LABORATORY TEST ABNORMALITIES

All laboratory test results captured as part of this study will be recorded following institutional procedures and stored in a database maintained by the DCC. Test results that constitute SAEs will be documented and reported.

The following laboratory abnormalities will be documented and reported appropriately:

a. Any laboratory test result that is clinically significant or meets the definition of an SAE
b. Any laboratory abnormality that required the subject to have study drug discontinued or interrupted
c. Any laboratory abnormality that required the subject to receive specific corrective therapy
d. Potential Drug Induced Liver Injury (DILI)
   Whenever possible, timely confirmation of initial liver-related laboratory abnormalities should occur prior to the reporting of a potential DILI event. All occurrences of potential DILIs meeting the defined criteria will be reported as SAEs. Potential Drug Induced Liver Injury is defined as:

   1. ALT or AST elevation > 3 times upper limit of normal (ULN) AND
   2. Total bilirubin > 2 times ULN, without initial findings of cholestasis (elevated serum alkaline phosphatase)
   3. No other immediately apparent possible causes of AST/ALT elevation and hyperbilirubinemia, including, but not limited to, viral hepatitis, pre-existing chronic or acute liver disease, or the administration of other drug(s) known to be hepatotoxic.

#### 8.9. PREGNANCY

If, following initiation of the investigational product, it is subsequently discovered that a study subject is pregnant or may have been pregnant at the time of investigational product exposure, including during at least 5 half lives (~ 49 days) after product administration, the investigational product will be permanently discontinued.

Investigators will immediately notify BMS Worldwide Safety: of the event using the Pregnancy Surveillance Form in accordance with SAE reporting procedures

Follow-up information regarding the course of the pregnancy, including perinatal and neonatal outcome and, where applicable, offspring information will be reported on the Pregnancy Surveillance Form.

Any pregnancy that occurs in a female partner of a male study participant should be reported to BMS. Information on the pregnancy will be collected on the Pregnancy Surveillance Form.

#### 8.10. OVERDOSE

An overdose is defined as the accidental or intentional administration of any dose of a product that is considered both excessive and medically important. All occurrences of overdose will be reported as an SAE.

#### 8.11. OTHER SAFETY CONSIDERATIONS

Any significant worsening noted during interim or final physical examinations, electrocardiograms, x-rays, and any other potential safety assessments, whether or not these procedures are required by the protocol, should also be recorded as a nonserious or serious AE, as appropriate and reported accordingly.

##### 8.12. DATA AND SAFETY MONITORING BOARD

The primary role of the Data and Safety Monitoring Board (**DSMB**) is to advise on scientific, safety, ethical and other policy issues relating to the study. DSMB members will:

a. Familiarize themselves with the study protocol, communicate by teleconference or email and develop a charter.
b. Propose appropriate analyses and meet periodically to review developing outcome and safety data.
c. Designate a board member to record and maintain minutes for all meetings.

The investigators will tabulate AEs and SAEs and report summary statistics of key outcomes to the DSMB at each meeting (every 6 months) to allow appropriate assessment of the relative risks and benefits.

#### 9. STATISTICAL CONSIDERATIONS

##### 9.1. SAMPLE SIZE CALCULATION

We designed this pilot study to assess the feasibility of conducting a large-scale phase III RCT and plan to enroll a relatively small number of patients over a limited period of time. As a feasibility study, we provide an assessment of the randomization rate to inform the design of the future trial. Some enrolled patients may not undergo transplantation during the study period. Other enrolled patients who undergo transplantation may not be eligible for randomization. We estimate that 60% of enrolled patients will be eligible for randomization. We plan to enroll 84 patients and estimate that 50 (84 * 0.6) will be eligible for randomization, and 40 (80%) will be randomized (50 * 0.8). We selected a sample size of 40 patients, as this will generate 95% confidence intervals of ±12% for the randomization rate. Thus, this sample size will allow us to estimate a randomization rate of 80% within a 95% confidence interval of ±12%.

We do not anticipate that this pilot study will detect a statistically significant difference in clinical outcomes (e.g., DSA development) between the 2 groups. However, this study will generate an estimate and 95% confidence intervals of the incidence of DSA development and other important clinical outcomes among lung transplant recipients treated with Belatacept-based immunosuppression. This will be important in making appropriate sample size calculations for the future phase III RCT.

##### 9.2. STATISTICAL ANALYSES

We will compare baseline characteristics across groups using t-tests (or Wilcoxon-Rank sum tests if the data are not normally distributed) and chi-square tests. We will use survival models (Kaplan-Meier method using the log rank test and Cox proportional hazards models) to compare freedom from DSA, CLAD, re-transplantation, and survival between the 2 groups to enhance the power of the analyses. We will use Cox proportional hazards models to adjust for covariates that include age, gender, race, primary diagnosis, HLA mismatch status, PGD grade, episodes of acute rejection, and CARV infection. Because of the relatively small sample size in this pilot study, we will adjust for only one covariate at a time and will use separate Cox models for each covariate. Thus, we do not plan to develop risk models for the outcome. However, in the larger pivotal trial, we would have an opportunity to use more complex statistical models. Similarly, we will analyze secondary outcomes using the Kaplan-Meier method and the log rank test and Cox models. All of the analyses will be conducted on an intention-to-treat basis that includes all subjects who are randomized. However, we will also conduct secondary analyses on a per protocol basis, recognizing that these can be potentially biased.

#### 10. STUDY ORGANIZATION

The study Steering Committee provides the leadership for the study group and has the decision-making authority for protocol changes. The Steering Committee consists of the 2 site PIs and the DCC Director (Drs. Huang, Hachem, and Schechtman). The DCC, directed by Dr. Schechtman, coordinates the implementation of the study and is responsible for developing data entry forms, electronic database, quality control systems, training personnel in the use of data entry systems, randomization procedures, analysis of data, providing study reports to the DSMB, the FDA, and other regulatory entities. The Clinical Coordinating Center, directed by Dr. Hachem, works closely with the DCC on study operations and is responsible for holding the FDA IND, maintaining and updating the study registration on ClinicalTrials.gov, and ensuring timely reporting of adverse events to the appropriate regulatory bodies.
